# Assessing the readiness of Oxford Nanopore sequencing for clinical genomics applications

**DOI:** 10.1101/2024.10.30.24316253

**Authors:** Judith Arres, Santosh Elavalli, Shalini Behl, Daniel Matias Sanchez, Ayesha Al Ali, Abdelrahman Ahmed Yehia Abdelaziz Saad, Azza Attia, Cyla Minas, Sharika Pariyachery, Shariq Ahmed, Fatmah Aldhuhoori, Nitu Thulasidharan, Gurunath Katagi, Omar Soliman, Shilp Purohit, Vinay Kusuma, Thyago Cardoso, Luis F Paulin, Philippe Sanio, Joseph Mafofo, Haiguo Wu, Val Zvereff, Albarah El-Khani, Fahed Al Marzooqi, Tiago R Magalhães, Fritz Sedlazeck, Javier Quilez

## Abstract

Long-read sequencing (LRS) technologies, namely Oxford Nanopore Technologies (ONT) and Pacific Biosciences, have emerged as promising solutions to overcome the limitations of short-read sequencing (SRS). Nevertheless, the still higher sequencing error rates compared to SRS, need for customized pipelines, rapidly updating software and incipient scalability are proving the adoption of the ONT for standard clinical practice to be challenging. Here we assess the performance of ONT (R9 and R10 chemistries) in comparison to Illumina and MGI across 17 well-characterized reference samples with 11 clinical variants representing 9 different genetic diseases. To enable this, we have implemented a production-ready pipeline including SNV, INDEL, STR, SV and CNV detection together with reporting key summary metrics to ensure high quality of data at production sequencing level. Our results show high accuracy of ONT across SNV (F-score >0.975) and SV, but still several weaknesses across INDEL (F-score<0.80). However, we highlight that ONT accurately detected all four pathogenic INDELs as well as the performance improvement in exons and with the newer R10 chemistry. We further demonstrated the importance of long-reads to detect clinical-impacting variants such as a *FMR1* pathogenic expansion, often misclassified by SRS as premutation range. Some issues remain as long-read analysis reported the wrong CNV genotype in one of the medical case samples. Remarkably, our multi-platform analysis and Sanger validation discovered a 1-bp error in the Coriell annotation for a cystic fibrosis causing INDEL in GM07829. Overall, this work highlights the readiness of ONT for clinical applications and large-scale operations.

## INTRODUCTION

The advent of long-read sequencing (LRS) was possible due to its increased read size from hundreds of base pairs (short reads) to multiple thousands or even millions of bases in one continuous read. These longer reads can resolve repetitive regions and improve the identification of structural variations (SV) (Mahmoud et al., 2024). This has enabled a more comprehensive insight into the diversity of the human genome, such as tandem repeats, centromeres and telomeres, which play critical roles in chromosome stability and aging processes (O’Sullivan & Karlseder, 2010). LRS further provides phasing information (i.e., determination of which alleles occur together on the same chromosome) that is crucial for understanding the inheritance of genetic traits accurately (Logsdon et al., 2020). Over the past decade, long reads have led to multiple novel insights across evolution, diversity of population and medical research.

Furthermore, we have learned more about certain limitations of short read sequencing (SRS) from incomplete assemblies, over the representation of repeats to the detection of certain genomic alterations such as SV. Nevertheless, SRS remains the work horse of genomics producing ever larger collections of genomes and exon data to study rare and complex diseases across the world. However, these studies might identify associated alleles with certain diseases but as often discussed fail to identify the causative allele due to lack of resolution or other reasons. This might be overcome by LRS in the near future as they increase the scalability by reduction of sequencing errors, costs and sample requirements.

The two main LRS technologies are Pacific Biosciences (PacBio) and Oxford Nanopore Technologies (ONT). Both are emerging as effective and potential solutions for clinical applications (reviewed in (Oehler et al., 2023). Given the rise of LRS multiple long-read population scale projects are under way or have been carried out (De Coster et al., 2021). More recently, All of Us, M42 and Genomics England have begun large scale sequencing based on ONT whole-genome sequencing (WGS) in the clinical genomics space. This is also partly motivated by multiple examples where LRS was beneficial to detect causative SV, such as significant deletions in the *DMD* gene leading to Duchenne muscular dystrophy (DMD) (Geng et al., 2023). LRS also excels in identifying repeat expansions in high GC content regions, areas where SRS often struggles, crucial for diagnosing disorders like Fragile X syndrome (FXS) (Stevanovski et al., 2022). Furthermore, ONT plays a critical role in variant phasing, essential for understanding inheritance patterns and pinpointing origins of de novo mutations, as shown by (Cretu Stancu et al., 2017). In addition, ONT’s capacity to differentiate clinically relevant genes from pseudogenes further enhances diagnostic precision, notably in identifying the *GBA* gene associated with Parkinson’s disease, reducing testing errors (Leija-Salazar et al., 2019). These examples have only been possible by the continuous development and overcoming challenges in applying LRS.

Specifically, ONT came a long way from being unreliable and suffering from 20%+ error rates to now being an established production platform. This transition is continuing with software releases that improve the error rate and establish even longer reads and methylation signals. These advancements demonstrate ONT’s superiority in identifying known causative alleles in regions of repeats compared to SRS. This is especially worth highlighting over certain pseudogenes or in general challenging medically relevant genes (CMRG). Furthermore, the speed by which ONT can operate makes it an encouraging instrument for rapid clinical implementation. Nevertheless, certain issues slow down the quicker adoption of ONT sequencing in genomics research and clinical applications.

Bioinformatic pipelines for ONT sequencing data often require specific optimizations, and dedicated callers are still being developed or evaluated (see **Discussion** and **Supplemental Note 1**). We believe it is important to integrate high-quality standards into ONT analysis pipelines to make ONT more scalable and production ready. Besides, homopolymer regions still inflate error rates across all platforms, with LRS being the most affected. These are often traceable, as ONT is one of the few technologies that directly sequences native DNA with all its advantages (i.e. methylation) but this also complicates noise patterns in its signal. Refining error models, such as those integrating machine learning to correct for homopolymer distortions, has shown promise in improving traceability (Wick et al., 2019). Additionally, more challenging benchmarks from Genome in a Bottle (GiaB) and other assembly derived variants showcase the ability of ONT to correctly identify mutations in these repetitive regions (Zook et al., 2016).

Most sequencing technologies are heavily tested on a relatively small number of well characterized reference samples such as the GiaB datasets (Krusche et al., 2019; Zook et al., 2016). Besides, many of the variant calling benchmarks have been based on such datasets too and have focused on single-nucleotide variants (SNV) and small insertions/deletions (INDEL). Therefore, it remains unclear how ONT and other sequencing technologies perform on independent datasets and other forms of genetic variation such as tandem repeats in which ONT may outperform SRS.

In this work, we focus on the clinical validation of ONT for genomics and its applicability in large population efforts aimed at non-ethnicity-biased variant prioritization. As part of this, we utilized an optimized analysis pipeline (https://gitlab.g42healthcare.ai/bix/health2.0_scripts/-/tree/main/ont/pipeline) for the comprehensive identification of genetic variations relevant to clinical genomics. This pipeline balances comprehensiveness and efficiency, ensuring fast turnaround times for both clinical cases and large-scale population studies. To establish ONT and our pipeline we assessed 17 well-characterized Coriell reference samples on ONT (both R9 and R10 chemistries) and compared their performance to two SRS technologies (Illumina and MGI). In contrast to GIAB and other benchmark samples, we were able to assess variant calling genome wide but more importantly the performance on pathogenic alleles. The latter is of utmost importance to demonstrate the actual utility of sequencing for clinical applications.

The value of this work stems from sequencing several Coriell reference samples to assess the performance of Illumina, MGI and ONT (R9 and R10) sequencing technologies. These samples have been selected to represent clinically relevant mutations, which was not done before to this extent. In addition, we have consolidated the pipeline for the analysis of ONT WGS data which calls clinically relevant variant types (SNV, INDEL, SV, CNV and STR) which, as a novelty, integrates important quality control (QC) steps. Altogether, these have allowed us to conduct an unprecedented benchmark of variant calling performance from ONT WGS relative to SRS.

## RESULTS

### Performant analysis pipeline for ONT WGS data

ONT is less mature than other sequencing platforms in the availability of streamlined end-to-end analysis pipelines which also include dedicated clinically relevant software tools (e.g. SMN1/2 caller). Therefore, a scalable and accurate workflow must integrate variant-calling methodologies along with upstream base-calling, alignment steps, and quality metrics to ensure high-quality results (**Supplementary Note S1**).

At M42 we have sequenced one of the largest ONT cohorts to date, enabling us to consolidate a comprehensive pipeline for the analysis of ONT WGS data (**Supplemental Figure S1**). This pipeline captures improvements we had made in our primary and secondary analysis pipeline for ONT WGS data plus the addition of callers for CNV and STR. We run concurrent real-time base-calling using the Nvidia A100 tower connected to the PromethION 48 ONT sequencing instrument and the latest Dorado base-caller (https://github.com/nanoporetech/dorado). Concurrent base-calling means that both the DNA sequence and 5mC methylation marks on the DNA are extracted from the Fast5 file generated by the sequencer. Real-time base-calling indicates that such step completes virtually by the time the entire sequencing run completes. We then perform in the cloud (a high-performance computing platform tailored for large-scale genomic data analysis) the mapping to the reference genome sequence using Sentieon-accelerated Minimap2 (https://www.sentieon.com/). Subsequently the variant calling is performed with dedicated tools for SNV/INDEL (Clair3, (Zheng et al., 2022), SV (Sniffles2, (Smolka et al., 2024)), CNV (Spectre [https://github.com/fritzsedlazeck/Spectre]) and STR (Straglr, (Chiu et al., 2021) (see **Methods** and **Supplemental Note 1**)**. It i**s not the objective of this work to present a ready-to-use pipeline we claim is superior to other possibly existing ones. That said, we have made the code available as a reference for others.

### A high-quality diverse set of 17 WGS reference samples sequenced on multiple HTS technologies

We identified 17 Coriell reference samples to assess the ability of high-throughput sequencing (HTS) technologies to accurately detect genetic variants. Coriell reference samples, sourced from the Coriell Institute for Medical Research, are well-characterized specimens for which biological material (e.g. cell lines or DNA) can be ordered and for which genetic “truth sets” exist and are available for the scientific community. For each of the 17 Coriell reference samples, we ordered cell lines (and not DNA) to avoid the fragmentation commonly observed in DNA, ensuring high-quality DNA for optimal long-read ONT sequencing (see **Methods**). We ordered cell lines for two types of Coriell reference samples. The first type included a parent-offspring trio (GM24143, GM24149 and GM24385) that has been extensively whole-genome sequenced by the scientific community, with publicly available truth sets of genetic variants (SNV and INDELS) across the genome (Zook et al., 2019) (**Supplemental Table S2**). The second type consisted of 14 samples derived from individuals affected by specific disease, for which the pathogenic genetic variants are well-documented (**Supplemental Table S2**). In each sample, we expect to detect the phenotype-causing genetic variant, which should be absent in the remaining samples (as they are known not to suffer the disease). We chose these specific Coriell reference samples to assess our accuracy to detect not only small variants – namely SNV and INDEL – but also other forms of genetic variation such as CNV deletions as well as STR.

We performed ∼30X WGS for each of the 17 Coriell cell lines on long-read ONT with R9 and R10 chemistries, and SRS technologies (Illumina and MGI). The R9 chemistry has been used since June 2016. It has been discontinued in July 2024 in favor of the new R10 chemistry. By including both ONT chemistries, we were able to estimate the differences of R10 relative to R9, a comparison which remains relatively unexplored outside GIAB (Ni et al., 2023). We performed WGS for two samples (GM27631 and GM03620) in replicates to assess intra- and inter-run variation. Altogether, the final dataset included 76 WGS samples.

We evaluated the quality of the WGS datasets. First, we confirmed that all 76 WGS samples passed the high-quality standards we defined (**Supplemental Table S1** and **Supplemental Table S3**). We selected those standards based on previous experience from UK Biobank (Sudlow et al., 2015), 1000 Genomes (1000 Genomes Project Consortium et al., 2015) and GATK Best Practices (DePristo et al., 2011). Overall, we generated more than 100 Gb of WGS data per sample across all three platforms (**Supplemental Figure S2** and **Supplemental Table S3**). The average genome coverage in ONT R9 samples (∼45X) was on average one third higher compared to the R10 samples (∼33X). This most likely mirrors the known lower productivity of R10 (Ni et al., 2023) that we confirmed also in our data (median yield was 103 and 141 Gb for R10 and R9, respectively) Additionally, the genome-wide coverage for ONT samples (∼40X) was generally higher than that of SRS samples (∼35X). However, despite the higher overall genome-wide coverage in ONT, the percentage of base pairs with >10X coverage was unexpectedly lower in ONT samples (<94%) compared to SRS samples (>95%), suggesting that ONT exhibits greater regional variability in coverage.

We also analyzed yield variation across the platforms. In Illumina and ONT R9 we observed the smallest range variation in yield across the 17 samples (93-131Gb and 122-163Gb, respectively). Variation in yield was notably higher for MGI (103-171Gb) and three times higher in ONT R10 (72-198Gb) compared to R9. In **Supplemental Table S3**, we can also see how read accuracy is still much lower in ONT compared to SRS, as previously reported (Amarasinghe et al., 2020). While we observed close to 90% of SRS sequencing bases with quality scores >Q30, a similar proportion of ONT sequencing bases only achieved >Q10. We could detect the improvement in read accuracy in R10 relative to R9 (median ∼86% and ∼80% bases >Q10, respectively). That improvement is consistent at increasing quality thresholds as well as when comparing average values (**Supplemental Figure S3**). In addition to increased read accuracy in R10, we also detected larger read lengths in that chemistry compared to R9 (**Supplemental Figure S4**). Although we found >95% of mapping rate across all platforms, both ONT chemistries remarkably achieved the highest (>99%). Given these very high mapping rates across samples, yield translated into the proportionally expected effective mapping coverage values.

In summary, compared to SRS we found that ONT (R9 and R10 consistently) has (i) lower read accuracy, (ii) more dispersed coverage values and (iii) higher mapping rate; between the two ONT chemistries, R10 seems to have (i) lower yield, (ii) improved read quality and (iii) longer reads.

In terms of the number of variants called per sample, we observed platform-specific differences (see **Supplemental Figure S5**). Coriell samples sequenced with Illumina generated approximately 5.0M variants per sample, of which around 4.0M were SNVs and 1.0M were INDELs. MGI produced slightly fewer variants, averaging 4.9M per sample, with approximately 4.0M SNVs and 0.9M INDELs. ONT R9 identified around 5.5M variants per sample, of which 4.5M were SNVs and 1.0M were INDELs. Finally, ONT R10 had the highest detection rate, detecting about 5.7M variants per sample, including 4.5M SNVs and 1.2M INDELs. These differences not only reflect the inherent characteristics of each sequencing platform but also highlight the tendency of long-read sequencing technologies to generate a higher rate of false positives, requiring stringent post-variant-calling filtering to ensure result accuracy.

Then, we applied principal component analysis (PCA) to the SNV/INDEL called across all samples for additional QC. PCA is a powerful tool for reducing information across hundreds of thousands of genetic markers into distinct sample similarity patterns. This allowed us to uncover any potential problematic samples or batch effects. In first place, the PCA reflected the ancestries of the 17 Coriell reference samples. The combination of the first two principal components (PC), explain 20.58% of the observed variance. As shown in **Supplemental Figure S6a,** the single African American sample and the South American family trio (left and bottom data points, respectively) are clearly separated from the remaining Caucasian-descent samples. Note that each individual is represented by 4 data points, corresponding to the sequencing performed across ONT platform (R9 and R10), Illumina and MGI. Closer evaluation further confirmed (**Supplemental Figure S6b**) that samples from the same individual clustered relatively together. Moreover, PC1 and PC2 also grouped samples from the same family. We observed that PC4 clearly distinguishes between SRS LRS, while the separation between ONT R9 and ONT R10 remains minimal (see **Supplemental Figure S7**). The effect of the sequencing technology reflected in PC4 accounted for 6.47% of the observed variation. While this shows the platform’s influence on genotype calls, it remains minor and arguably has no impact on the subsequent analyses. Additionally, no outliers were observed in any of the PCs, indicating the absence of problematic samples. Overall, our results confirm the robustness of the dataset, ensuring the reliability of downstream analyses.

### Performance of SNV, INDEL and SV detection across the genome

We used the parent-offspring trio (GM24143, GM24149, and GM24385) to assess our SNV/INDEL variant calling performance genome-wide (**Supplemental Table S2**). We compared the SNV/INDEL call sets generated for each of their ONT (both R9 and R10), Illumina and MGI WGS samples against the publicly available golden-truth variants to calculate standard performance metrics such as recall, precision and F-score (see **Methods**). We investigated those performance metrics genome-wide as well as in exonic regions and CMRG. In all comparisons, we achieved very homogeneous metrics values across samples within each sequencing technology (as shown by the narrow interquartile ranges in all the boxplots in **Figure 1**), suggesting high consistency of the reported performance metrics.

**Figure 1.**
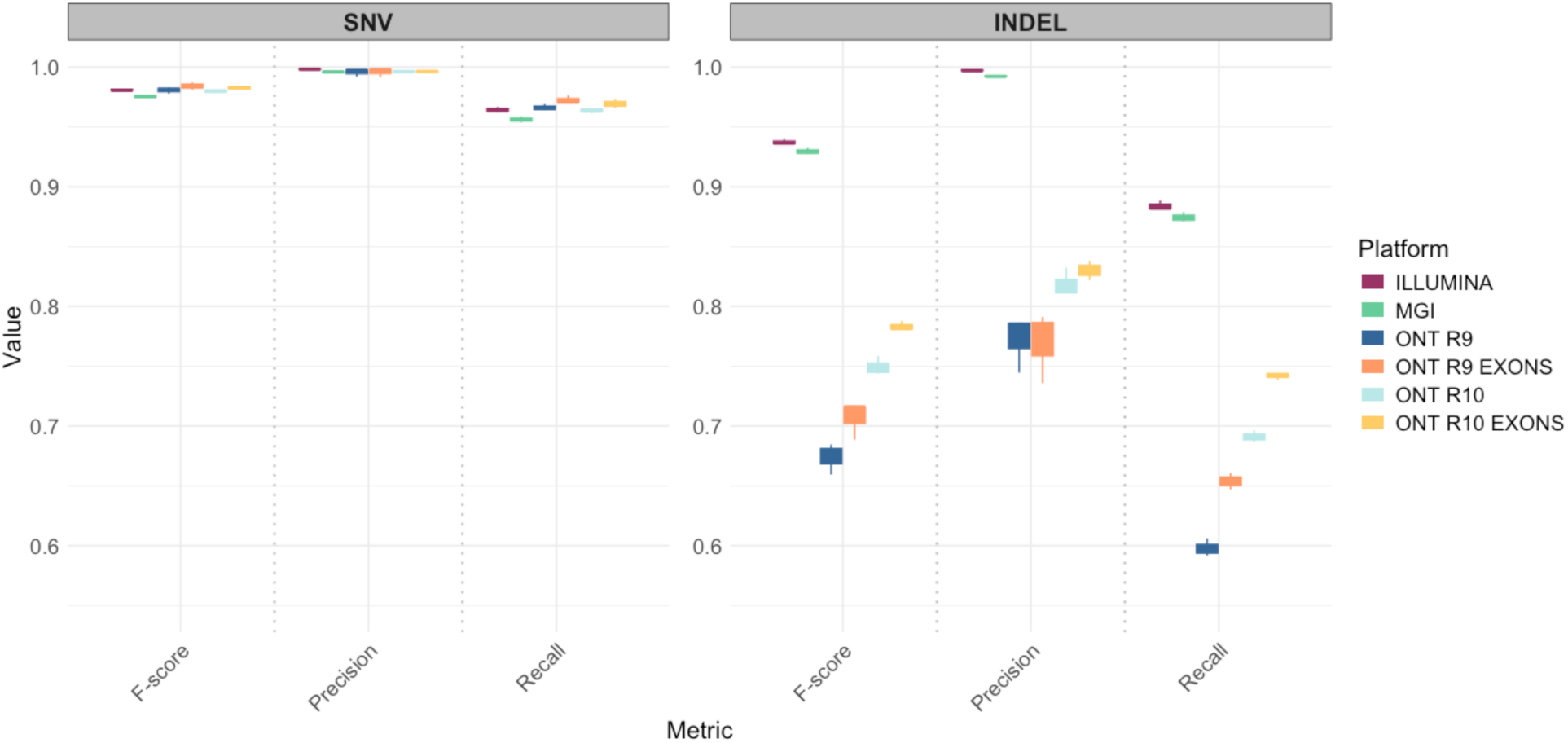
SNV and INDEL calling performance across sequencing platforms. F-score, precision and recall distribution for SNVs (left) and INDELs (right) across the different sequencing platforms. For Illumina, MGI and ONT (both R9 and R10), displayed are performance metrics in high-confidence regions genome wide. In addition, for ONT R9 and R10 shown are also performance metrics in exonic regions. Each boxplot encapsulates the corresponding metrics for the three Coriell samples with truth sets available (GM24143, GM24149, and GM24385).

In all three sequencing platforms we achieved high performance in calling SNV genome wide, as denoted by F-score >0.975 in any of the samples analyzed (**Figure 1**, **Supplemental Figure S8 and Table 1a**). As expected, we observed higher SNV precision (median = 0.997) than recall (median = 0.963) regardless of the sequencing platform. While Illumina exhibited slightly higher SNV precision compared to the other platforms, ONT (both R9 and R10) performed better than SRS in SNV recall, especially compared to MGI. Regardless, these values consistently support our capability to detect SNV in all platforms.

**Table 1.** Summary of genome-wide SNV/INDEL calling performance and detection of positive controls across sequencing platforms. (a) F-score performance metric values for SNV and INDEL calls separately for each of the samples in the Coriell trio with truth sets available. **(b)** Detection (✓) or not (✘) of the expected genotype for each of the 14 positive control Coriell reference samples. *For GM27631 and GM27632, detection of the precise genotype is absence of the MRD40-causing de novo present in their child (GM27630). Abbreviations: BRCA1 = Breast Cancer Type 1; MRD40 = Mental Retardation Autosomal Dominant 40; USH1C = Usher Syndrome Type IC; CF = Cystic Fibrosis; DMD = Muscular Dystrophy, Duchenne Type; SMA1 = Spinal Muscular Atrophy I; DM = Dystrophia Myotonica; FMR1 = Fragile X Syndrome; HD = Huntington Disease.

| Sample | Description | Variant Type | Illumina | MGI | ONT (R9) | ONT (R10) |
| --- | --- | --- | --- | --- | --- | --- |
| a |  |  | F-score |  |  |  |
| GM24143 | Trio (Mother) | SNV | 0.980 | 0.975 | 0.981 | 0.978 |
|  |  | INDEL | 0.936 | 0.928 | 0.678 | 0.744 |
| GM24149 | Trio (Father) | SNV | 0.981 | 0.975 | 0.978 | 0.980 |
|  |  | INDEL | 0.936 | 0.929 | 0.659 | 0.758 |
| GM24385 | Trio (Child) | SNV | 0.982 | 0.977 | 0.983 | 0.981 |
|  |  | INDEL | 0.940 | 0.932 | 0.685 | 0.746 |
| b |  |  |  |  |  |  |
| GM13708 | BRCA1 | SNV | ✓ | ✓ | ✓ | ✓ |
| GM27630 | MRD40 | SNV | ✓ | ✓ | ✓ | ✓ |
| GM27631* | MRD40 | SNV | ✓ | ✓ | ✓ | ✓ |
| GM27632* | MRD40 | SNV | ✓ | ✓ | ✓ | ✓ |
| GM10354 | USH1C | SNV | ✓ | ✓ | ✓ | ✓ |
| GM07828 | CF | INDEL | ✓ | ✓ | ✓ | ✓ |
| GM07829 | CF | INDEL | ✓ | ✓ | ✓ | ✓ |
| GM07830 | CF | INDEL | ✓ | ✓ | ✓ | ✓ |
| GM08211 | CF | INDEL | ✓ | ✓ | ✓ | ✓ |
| GM04099 | DMD | CNV (Deletion) | ✓ | ✓ | ✓ | ✗ |
| GM10684 | SMA1 | CNV (Deletion) | ✓ | ✓ | ✓ | ✓ |
| GM03990 | DM | STR | ✓ | ✓ | ✓ | ✓ |
| GM09145 | FXS | STR | ✗ | ✗ | ✓ | ✓ |
| GM03620 | HD | STR | ✓ | ✓ | ✓ | ✓ |

Accurately detecting INDELs is more challenging compared to SNV (i.e. small deletions or insertions 50–100 bp compared to single-point sequence changes). We confirmed that INDEL performance is worse for SNV, with lower precision and worse recall (**Figure 1** and **Table 1a**). We found that drop in the INDEL performance is very pronounced in ONT, with an F-score <0.80, precision <0.85 and recall <0.70 in all samples. In contrast, the same metrics exceeded >0.85 in SRS samples and were consistently higher in Illumina compared to MGI. While ONT performed worse than Illumina and MGI, we observed a substantial improvement in INDELs detection with the newer chemistry compared to the previous R9.

Clinical genomics applications utilize genetic variants with a functional impact. In practice, this typically translates into restricting analyses to coding regions of the genome, namely, the exons. Changes in exons have more impact and interpretable functional change, leading to phenotypic manifestations and, in many cases, clinical symptoms. In contrast, the functional impact of non-coding regions is less clear, causing that less attention is paid to those. Given that, we wondered whether SNV/INDEL variant calling performance using ONT sequencing data is higher in exons compared to the genome-wide results we reported above – this would increase the confidence in utilizing this sequencing technology in clinical applications. We found that the performance of ONT to call INDEL was higher in exons compared to genome wide (**Figure 1**). The largest improvement was in detecting true INDEL exonic loci, with 5.6% and 5.1% increases in the recall metric in R9 and R10, respectively. R10 also outperformed R9 in exons in terms of INDEL recall. Overall, this improvement in recall is particularly important, as it reduces the proportion of missed true variants or False Negatives (FN), which was more pronounced at the genome-wide level. Restricting our analysis to exons did not alter INDEL precision in R9 but it slightly increased that metric in R10. This means that while ONT may miss some true variants, those that are detected are generally accurate, reflecting a low rate of false positives (FP). On the other side, focusing on exons had little impact in the already high SNV calling performance we reported above (**Figure 1**). While recall for SNV slightly increased for those located in exons, SNV precision remained consistently high across in R9 and R10.

In the previous analysis we focused on exonic regions as these are more commonly used in clinical genomics applications. For similar reasons, other previous benchmarks have focused on genes that are medically important such as the panel of 73 actionable genes released by the American College of Medical Genetics and Genomics (ACMG) (Mahmoud et al., 2024; Mandelker et al., 2016; Miller et al., 2021). Others have particularly focused on medically relevant genes that are hard to impossible to characterize by SRS to highlight the potential of LRS to resolve these loci (Mahmoud et al., 2024; Mandelker et al., 2016; Miller et al., 2021). Likewise, here we evaluated the performance of the three sequencing technologies in a previously defined set of 273 CMRG using GM24385, for which truth values in these regions exist. In line with the genome- and exome-wide performance results (**Figure 1**), ONT (both R9 and R10) performed as well as Illumina and even better than MGI when calling SNV (**Supplemental Figure S9**). While precision was slightly lower for ONT compared to the two SRS technologies, recall was clearly higher for LRS even compared to Illumina. As previously reported (Mahmoud et al., 2024; Wagner et al., 2022), detecting INDEL within CMRG is particularly challenging for SRS, as denoted by the decrease of 0.07 in precision relative to the genome-wide performance. In contrast, the impact in ONT is lower with only a drop of 0.03 in precision. Again, the improvement, especially in recall, of R10 relative to its predecessor R9 is also noticeable in CMRG.

We also used the well-characterized GM24385 sample to compare SV between LRS and SRS, for which SV truth sets exist. We observed high precision (>0.90) in ONT and Illumina genome-wide (**Supplemental Table S4a**). On the other side, recall in ONT was more than two times higher than in Illumina (>0.60 and 0.26, respectively), which translated into an aggregated F1 score in ONT almost twice (∼0.75) as high as Illumina’s (0.40). This higher SV performance of ONT was even more pronounced in CMRG (**Supplemental Table S4b**). Precision for both Illumina and ONT, along with Illumina’s recall, exhibited consistency with genome-wide performance (**Supplemental Table S5b**). In contrast, ONT’s recall and F1 values boosted close to 0.90. All SV performance metrics were very similar between ONT’s R9 and R10.

### Performance detection of disease-causing mutations

We extended the evaluation of the performance of the three sequencing technologies to detect disease-causing mutations in 14 Coriell reference samples. These samples contained 11 mutations (3 SNV, 3 INDEL, 2 CNV and 3 STR) linked to 9 different diseases of high clinical relevance – e.g. breast cancer (BRCA), cystic fibrosis (CF) or DMD (**Supplemental Table S2**). In each of these samples, we assessed the presence or absence of the associated known mutation, for which genomic coordinates and changes in the genome reference sequence are publicly available.

Both SRS and LRS precisely detected the expected three disease-causing SNV evaluated (**Table 1b** and **Supplemental Table S5**). In GM27630, mental retardation autosomal dominant (MRD40) is caused by a de novo c.2127T>G (p.Tyr709*) pathogenic dominant mutation which should therefore be absent in the two parents, also included in our dataset (GM27631 and GM27632). By analyzing the trio samples from each sequencing platform, we confirmed as expected a single copy of the pathogenic variant in the proband and its absence in both parents.

We evaluated three INDELs causing CF (556delA, del508 and 557delT) – delF508 present in three of the Coriell samples and one of them (GM07830) bearing two of the CF-associated mutations (Supplemental Table S2). We confidently detected the three phenotype-causing INDEL in all 4 sequencing technologies/chemistries. Interestingly, our multi-platform high-quality sequencing approach detected a likely 1-bp annotation error in the GM07829 Coriell reference sample. According to the Coriell Institute for Medical Research, the CF phenotype in this sample is caused by a deletion of an adenine (A) relative to the genome reference sequence at the position chr7: 117,531,049 bp (Zielenski et al., 1991). Instead, we detected a deletion of the immediately downstream thymine (T) in all 4 WGS samples for GM07829 (chr7: 117,531,050 bp) (Supplemental Table S5). We ruled out mapping or variant calling artifacts as well as confirmed the T deletion through visual inspection in IGV (Figure 2a–d). Moreover, through Sanger sequencing we confirmed the T deletion at chr7: 117,531,050 bp (Figure 2e) and thus orthogonally validated the genotypes predicted by all HTS technologies. Of note, the INDEL we predict in all our samples has the same functional impact on the CFTR protein which leads to the CF phenotype presented by the individual sourcing GM07829.

**Figure 2.**
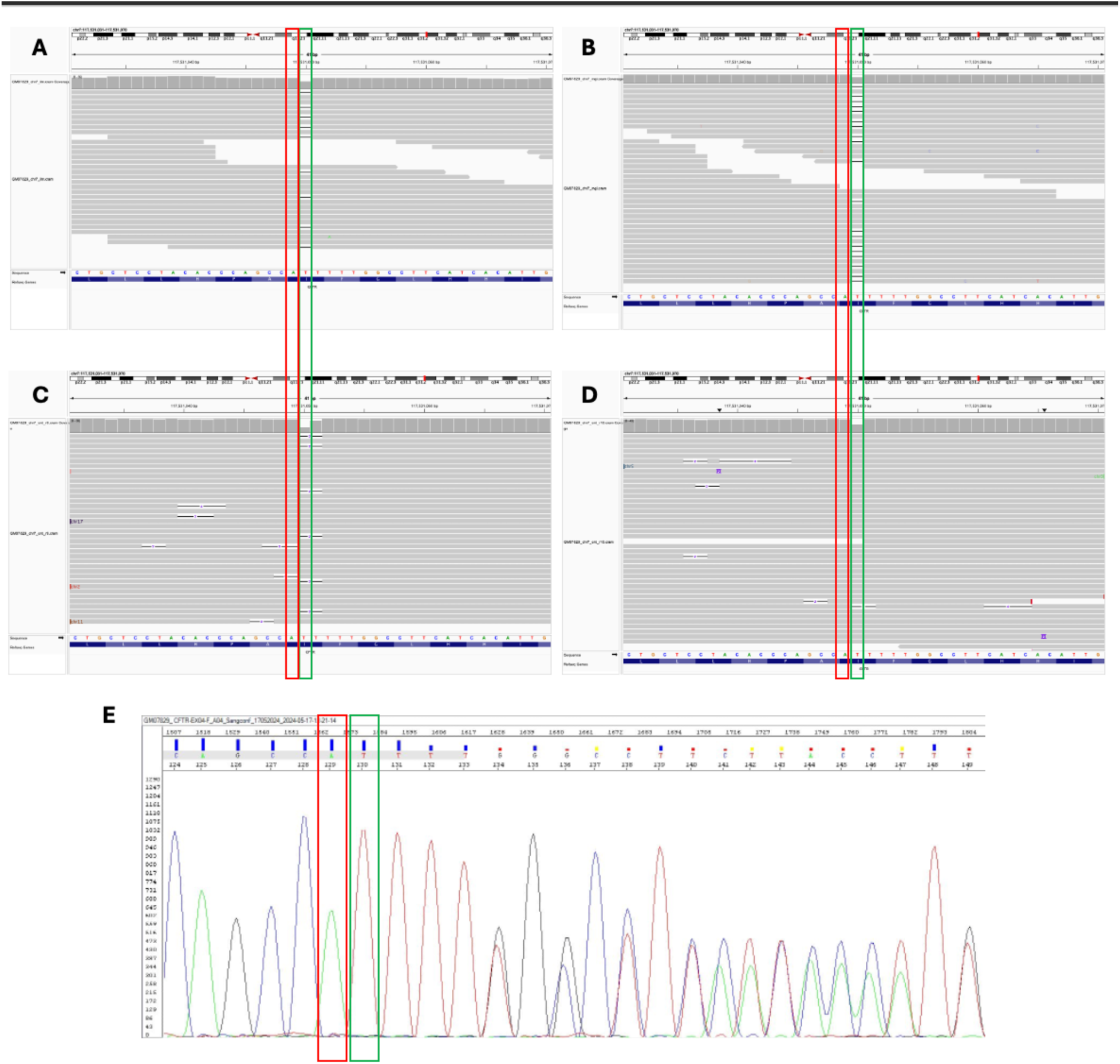
Across-platforms HTS and Sanger orthogonal validation uncover a 1-bp annotation error in GM07829. **(a–d)** IGV snapshots supporting the absence of the 55delA INDEL (chr7: 117,531,049 bp) annotated in Coriell for GM07829 but instead a 1-bp downstream T deletion (chr7: 117,531,050 bp) with the same detrimental functional effect. **(e)** Orthogonal validation of the HTS results through Sanger sequencing.

We accurately genotyped the CNV responsible for DMD in GM04099. Both SRS and ONT R9 detected a single copy (i.e., heterozygous) of the deletion of exons 49–52 in the *DMD* gene. However, in the ONT R10 sample, the Spectre CNV caller in our pipeline failed to identify this pathogenic deletion (**Supplemental Table S5**). The sequencing read alignment profiles for GM04099 in ONT R10, however, remain consistent with the expected heterozygous genotype. Specifically, the mapping coverage in the flanking regions of the CNV locus aligned with the genome-wide average (∼30X for ONT R10), with approximately half the coverage observed within the deletion region (**Supplemental Figure S10**), thus ruling out the influence of abnormal coverage on the genotype call. Sniffles2 caller was also evaluated, though its results were less precise. Altogether, we believe these findings point to limitations of ONT-dedicated variant calling algorithms, rather than an issue with the sequencing of this region with ONT.

Furthermore, the deletion of exons 7 and 8 in the *SMN1* gene illustrates how ONT is catching up in sequencing accuracy and software developments with SRS. This variant is particularly relevant in the clinical context due to its association with spinal muscular atrophy (SMA), a severe genetic disorder. SMA is caused in most cases by deletions in the *SMN1* gene, but accurately genotyping these deletions is complicated by the presence of the nearly identical paralog SMN2. Illumina developed a dedicated caller to resolve genetic variation in *SMN1* and *SMN2*, enabling accurate screening of pathogenic deletions in *SMN1* – this caller is integrated into Illumina’s DRAGEN. We implemented its open-source version (X. Chen et al., 2020) for MGI. Additionally, we introduced an alpha version of an *SMN1* and *SMN2* caller, named *Sillago*, specifically developed by ONT (Oxford Nanopore Technologies, 2024). As expected, across all sequencing technologies evaluated, we correctly genotyped the SMA GM10684 sample as homozygous for a deletion of exons 7 and 8 in *SMN1*, while identifying two normal copies of *SMN2* (**Supplemental Table S5**). Notably, the novel *Sillago* caller accurately resolves this challenging locus in both R9 and R10.Our evaluation also included tri-nucleotide expansions leading to disease phenotypes (**Supplemental Table S2**), using specific STR callers in our analysis pipeline. Expansion Hunter (Dolzhenko et al., 2017) was employed for Illumina and MGI, while Straglr (Chiu et al., 2021) was applied for ONT data. In our study, we included a sample affected by dystrophia myotonica (DM), a genetic disorder caused by the expansion of more than 50 CTG repeats in the *DMPK* gene. Specifically, the GM03990 sample was reported to have between 50 and 80 CTG repeats. In our analysis, we observed 64, 57, 78, and 79 copies using Illumina, MGI, ONT R9, and ONT R10, respectively (see **Figure 3)**. Similarly, Huntington disease (HD) is caused by >36 copies of the CAG tandem repeat in the *HTT* gene – the GM03620 sample in our study is expected to present 60 copies of such repeat. Illumina detected 55 repeats, MGI identified 68, while ONT R9 and R10 found 78 and 79 copies (see **Figure 3)**. Altogether, for both GM03990 and GM03620, in all sequencing technologies we predicted the STR copy number in the pathogenic range for DM and HD, respectively (**Supplemental Table S5**).

**Figure 3.**
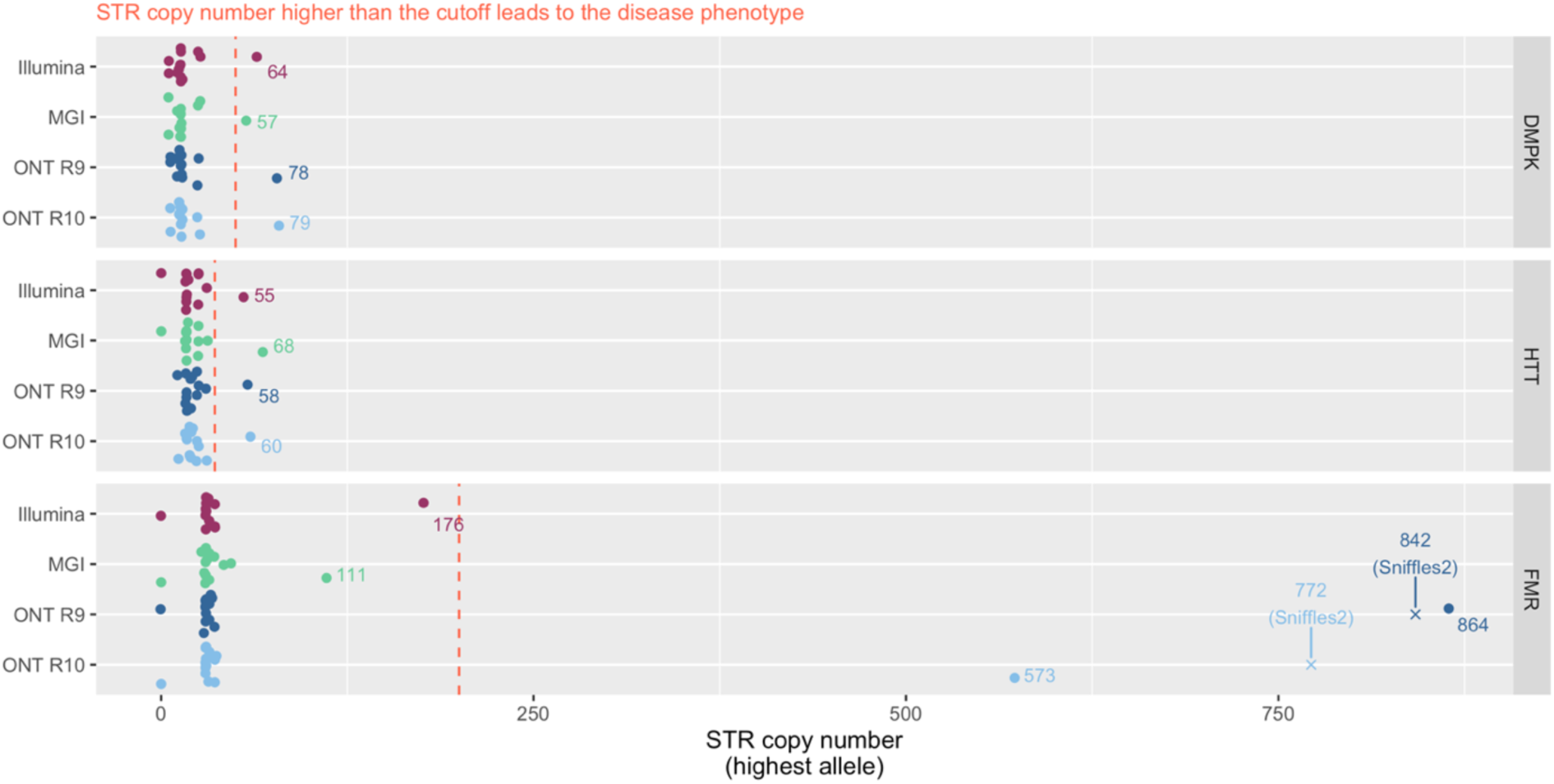
Detection of STR expansions across sequencing platforms for genes FMR1, HTT, and DMPK in the 14 Coriell Samples. The first panel shows the detection of CTG repeats in the DMPK gene where expansions over 50 repeats are associated with myotonic dystrophy. The intermedium panel displays the detection of CAG repeats in the HTT gene, with expansions beyond 36 repeats, characteristic of Huntington’s disease. And finally, the last panel illustrates the detection of CGG repeats in the FMR1 gene, where expansions exceeding 200 repeats are indicative of Fragile X syndrome.

The third evaluated STR was a CGG expansion in the *FMR1* gene which causes FXS when present at >200 copies (Hagerman et al., 2017) (**Supplemental Table S2**). SRS failed to precisely call the pathogenic form of this STR in the GM09145 included in our study. In Illumina and MGI we detected, respectively, 176 and 111 copies of the CGG-repeat (**Figure 3** and **Supplemental Table S5**), which fall below the >200 value defining the expected disease phenotype. This demonstrates the limitations inherent to SRS platforms in accurately detecting STR, mainly due to average read length and the extensive size of these specific repeat sequences. In contrast, we consistently detected >200 copies of this CGG repeat linked to FXS in the sequencing of GM09145 with ONT R9 or R10 (**Figure 3**). We noted considerable variation between R9 and R10 chemistries (864 and 573 copies, respectively) (**Figure 3**).

Because the FXS-linked CGG repeat is relatively long, we hypothesized that it could be detected too by the Sniffles2 (Smolka et al., 2024) SV caller in our ONT pipeline (which is meant to target larger-scale structural variation events). Indeed, we confirmed that Sniffles2 detected the CGG-repeat within the *FMR1* gene as an CGG-based insertion relatively matching the length detected by Straglr, with 842 repeats for ONT R9 and 772 for ONT R10 (**Figure 3**). This approach using different callers for variant detection, highlighted the robustness of LRS in identifying variants confidently.

In summary, we correctly identified the expected genotype in 12 out of the 14 (>85%) Coriell positive control samples across all four sequencing platforms (Illumina, MGI, ONT R9, and ONT R10) (**Table 1**) – in other words, overall we precisely detected the expected genotype in 53 out of all the 56 (95%) Coriell positive controls across sequencing platforms. Each of the 11 disease-causing mutations we evaluated (**Supplemental Table S2**) is only expected to be found in the associated Coriell positive control, but not in the others (which can then be treated as true negative controls). Indeed, we verified that this was consistently in all the cases (**Supplemental Table S5**), which translates into a genotyping accuracy of nearly 100%.

Finally, for ONT only we tested the consistency within (intra) and between (inter) sequencing runs using two of the Coriell positive controls: GM27631 (carrier of c.2127T>G in MRD40) and GM03620 (CAG repeat in *HTT*). Intra-run tests examine the uniformity and stability of sequencing results within a single batch or run, detecting potential variability that may occur during the sequencing process. Inter-run, on the other hand, extends this quality assessment across multiple sequencing runs, which can occur on different days, involve different reagents or use different machines, ensuring robust reproducibility across different conditions. In the intra-run tests, we sequenced each of the two samples twice within the same run. The c.2127T>G mutation in GM27631 was successfully detected in both intra-run replicates for both R9 and R10. Similarly, the predicted CAG repeat numbers in GM03620 were consistent between the intra-run replicates for R9 (58 and 58 repeats) and R10 (59 and 60 repeats). For the inter-run analysis, we sequenced the same two samples in two separate sequencing runs. The c.2127T>G mutation was detected in all inter-run replicates, and the STR repeat numbers were again consistent across runs (R9: 58 and 58; R10: 59 and 60). Altogether, our results demonstrated satisfactory intra- and inter-run replicability of ONT sequencing.

### Across-platforms concordance

In addition to the across-platforms differences relative to reference truth sets, we further explored systematic differences in variant calling between platforms. We noticed that ONT, both R9 and R10, call an excess of SNP/INDEL around the centromeres compared to other parts of the chromosome (**Supplemental Figure S11**). That is also visible for SRS to less extent and only in certain chromosomes. ONT also shows a higher proportion of variants in the region of chromosome 6 which harbors the human leukocyte antigen (HLA). These observations are likely driven by inaccuracies in the GRCh38 reference genome, particularly in highly repetitive and structurally complex regions, which result in misaligned reads. While SRS platforms typically discard these reads based on stringent mapping quality thresholds, long-read technologies retain a larger proportion of these misaligned reads, potentially contributing to the observed increase in variant calls.

We investigated genotype concordance between sequencing platforms. For SNV and INDEL, we measured concordance as the Jaccard index value calculated from the call sets in any two pair of samples compared (see **Supplemental Figure S12**) – we calculated the Jaccard index only for pairs of sequencing samples derived from the same Coriell specimen and for SNV and INDEL separately (see **Methods**). In SNV we detected >78.5% genotype sharing between any platform and chemistry compared (median = 82.4%, max = 92.0%) – we evaluated ∼4 million sites genome wide. The two SRS technologies showed the highest concordance (>90%) followed by the ∼87% similarity between ONT chemistries (**Supplemental Figure 12** and **Supplemental Table S6**). The overall concordance between LRS and SRS was the lowest at ∼80%. Similarity between ONT and Illumina was higher compared to ONT and MGI. Unexpected similarity values between the newer ONT chemistry R10 and SRS was lower compared to R9 and SRS. As expected, similarity patterns in INDELs were lower across all platform comparisons (median = 59.9%, range = 50.8–82.6%). Intra-SRS comparisons still showed the highest similarity (81.2%) but ∼10% lower compared to SNV (90.6%). Any ONT-based comparison fell below 70% similarity, even the R9 versus R10, which for SNV was >86% in any sample. Contrary to the SNV results—where R9 demonstrated better concordance with SRS than R10—the R10-SRS comparison for INDELs exhibited higher similarity (>61.7%) relative to the R9-SRS comparison (∼55.2%). Altogether, these results reinforce that accurate calling of INDEL is more challenging than for SNV, especially for ONT.

## DISCUSSION

In this work, we evaluated (and contributed to) the readiness of ONT for clinical genomics applications and population studies. Improvements in this LRS technology and its advantages over SRS have resulted in increasing clinical and research work leveraging ONT (Mahmoud et al., 2024; Oehler et al., 2023; Wick et al., 2019).

Yet, some questions remain and slow down a faster adoption of ONT. Is the lower read accuracy of ONT relative to SRS still translating into less trustable variant calls? Is the analysis of ONT production-ready for fast turnaround and / or large volumes of samples and able to target important genetic variants?

To address these questions, we first have compared the performance of ONT to Illumina and MGI across 17 well characterized samples including pathogenic SNV, INDEL, CNV and STR. Most importantly is the analysis of the 14 samples carrying pathogenic alleles which have not been previously assessed across these technologies. We found that the performance of ONT is catching up with SRS, although differences across the sequencing technologies in terms of detection biases persist. Our results showed that ONT is as good as SRS in detecting SNV and outperformed short reads in large and repetitive variants such as SV and STR. ONT’s weakest point is still at detecting INDEL (Amarasinghe et al., 2020) but it improves with the new R10 chemistry and in loci that are medically relevant and / or challenging for SRS. To further promote the integration of ONT, we leveraged these samples to establish a novel analysis pipeline that is capable of scaling and reporting all variant types. Importantly, the pipeline includes the most advanced and performing software for ONT as well as integrates important QC metrics to monitor and assess the quality of the samples being processed in production setups. We were able to establish this pipeline across all samples and show its utility across R9 and R10 data.

One important milestone missing in genomics and genetics is the simultaneous ascertainment of all variant types / classes together. Many association or general genetic studies remain focused only on either SNV/INDELs or other forms of variation (e.g. STR) and how these separately impact on certain phenotypes. Yet, all different variant types co-occur on the same DNA molecules and thus have equal opportunity to impact phenotypes. To illustrate this, despite the number of SV being only a small fraction compared to SNV and INDELS (roughly 23,000 SV versus 4–5 million SNV/INDEL), the SV typically impact a higher number of nucleotides along the genome. The primary reason we continue to study SNVs is our advanced knowledge and annotation methods for estimating their impact, especially when compared to SVs and STRs. This is in part because of detection biases and challenges of these more complex variants compared to short-read based SNV detection approaches. We must improve our detection of SVs and STRs on a population scale, which enables us to derive improved functional models for these complex variant types. This will require conducting LRS at scale as it has begun in certain consortia. To promote this development, we presented our state-of-the-art pipeline that includes SNV, INDEL, STR, SV and CNV calling together with the targeted *SMN1* and *SMN2* caller. We included the assessment of multiple important quality metrics in the analysis pipeline to automate and scale discerning between high-quality and failed samples. This pipeline enabled us to investigate the performance of ONT R9 and R10 and the improvements in variant calling. We further achieved higher scalability by choosing speed-optimized software for ONT (e.g. Sentieon’s Minimap implementation) as well being early adopters of upgrades rolled out by ONT – for instance, replacing the Nvidia’s V100 connected to the PromethION sequencer by the more powerful A100 coupled with Dorado base-calling (instead of Guppy) enabled real-time base-calling, streamlined our ONT analysis workflow and reduced storage costs of large Fast5 files (∼700 GB per ∼30X human WGS). In this work, we have demonstrated the accuracy of the pipeline across multiple samples and demonstrated its ability to call all variant types together. This directly promotes the simultaneous assessment of all variant types at scale, which we believe is important over the next decade of genomics and genetics.

There are multiple advantages but also some disadvantages remain when implementing ONT sequencing. While we identified that the SNV accuracy of ONT is on par with the short read approaches: ONT (F1 = 0.978), Illumina (F1 = 0.980), MGI (F1 = 0.975). We still observed some deficiencies in the INDEL calling for ONT, especially in repetitive regions. This was also reported previously and seems to improve over the different generations of base-caller and variant callers (Logsdon et al., 2020; Wenger et al., 2019). We demonstrated a clear improvement in our work between the R9 and R10 chemistry where we measured a 5–7% increase in accuracy. Still, Illumina and MGI both show a higher accuracy for INDELs e.g. for GM24385 as 94.0% and 93.2%, respectively. Nevertheless, it is noteworthy that ONT identified correctly the presence or absence of all four pathogenic INDELs across the 14 samples. This indicates that the lower INDEL detection performance of ONT genome wide actually did not lead to a detection bias of pathogenic INDELS itself. This is an important observation as arguably these are the most important variants to identify. The improved detection of causative INDELs might be impacted by the occurrence of these variants in coding regions that are typically not part of tandem repeats or specifically homopolymers. We and others could demonstrate that in coding regions themselves the ONT INDEL accuracy is much improved compared to other repeat regions (Logsdon et al., 2020; Mahmoud et al., 2024). The detection of variants inside of tandem repeats further is actually boosted from ONT by the improved characterization of STR and SV themselves compared to Illumina or MGI. Over the past decade we learned much more about SV and STR and the impact of these alleles on human diseases and phenotypes (Fotsing et al., 2019; Liu et al., 2022). In our work, we demonstrated that SRS technologies can fail to accurately detect the full extent of repeat expansions in some cases, leading to the misclassification of pathogenic STR. This is clear for the sample GM09145, which carries a full mutation of the tandem repeat (>200 copies) impacting *FMR1* over silencing the gene and causing FXS. This sample was wrongly called by Illumina and MGI with both technologies reporting a pre-mutation range (50-200 repeats), which would not cause the diseases phenotype itself. This sample highlights the need and importance to utilize pathogenic variants for benchmarking to further improve our understanding about these important genomic variants.

In summary, our work shows that ONT identifies SNV genome wide as accurately as SRS. While the Achilles heel of ONT continues to be INDEL detection, this is less so in exons and in regions that are challenging for SRS whatsoever, plus we showed the improvement in the new R10 ONT chemistry. As a matter of fact, ONT accurately detected all four disease-causing INDEL here evaluated. Indeed, ONT performed similarly well as SRS in detecting other disease-causing variants we interrogated. Altogether, our results will provide guidance for organizations aspiring to incorporate ONT into their clinical workflows. To additionally help that process, here we share practical advice for the implementation bioinformatics pipelines for the analysis of ONT WGS data.

## METHODS

### Coriell reference samples

Lymphoblast cell cultures were purchased from Coriell Cell Repositories (Camden, NJ) and processed as per the supplier’s instructions. It was chosen to purchase direct cell lines and extract DNA in house rather than the DNA reference material to preserve the native form of DNA. Transportation of DNA may result into fragmentation which may have otherwise affected the long reads required for sequencing on the Oxford Nanopore platform. Upon receipt of cells in T-12.5ml tissue culture flasks, they were incubated overnight at 37*0*C. The following day, cells from the culture flasks were transferred to 50 ml centrifuge tube and centrifuged for 10 mins at 100Xg. The supernatant was discarded and the cell pellet was resuspended with cell culture complete medium (RPMI 1640 with 15% FBS). The cells were then evenly distributed (Approx. 1×10^6^) into 25 ml tissue culture flasks containing 10 ml of media and incubated at 37*0*C with 5% CO2. When adequate growth was seen, the cells were harvested and the number of cells was determined.

### DNA extraction

DNA extraction was done using the PureLink™ Genomic DNA Mini Kit following the manufacture’s protocol. Briefly the sample tubes were centrifuged at a maximum speed of 15,000 rpm for 5 minutes followed by the removal of growth medium without disturbing the cell pellet. The cells were resuspended in 200 µl of Phosphate buffer saline (PBS) solution and 20 µl each of proteinase K and RNase A were added to the tubes. This was followed by addition of 200 µl PureLink Genomic Lysis/Binding Buffer. The solution was mixed well by vortexing to obtain a homogeneous mixture which was then incubated at 55°C for 10 minutes. 200 µl of 96-100% ethanol was added to the lysate. Approximately, 640 µl of this lysate was transferred to a spin column and centrifuged for a minute. Collection tube with the supernatant was discarded and the spin column was placed into a new collection tube. The column was then washed twice with 500 µl of Wash buffer 1 and Wash buffer2 respectively. Finally, 200 µl of elution buffer was added to the column and centrifuged at maximum speed for 1 minute at room temperature for optimal yields of elute.

### Sequencing

#### Illumina

Whole genome sequencing (30X) library preparation was performed by using Illumina PCR free prep library kit. Following instructions from the manufacturer, gDNA input of 250 to 750ng was fragmented by Bead-linked transposome and ligation was done using IDT® for Illumina® UMI DNA/RNA UD Indexes Set A (96 Indexes, 96 Samples). All 24 samples were pooled on the basis of indexes compatibility and sequenced using the NovaSeq 6000 S4 Reagent Kit v1.5 (300 cycles) on the NovaSeq 6000 System.

#### MGI

Library preparation was done using MGIEasy PCR-Free DNA Library Prep (96 RXN) kit. A total of 900 ng DNA in 48 ul was used. Preparation steps included fragmentation, size selection, end repair, adapter ligation, denaturation, circularization and exo-digestion. Double-size selection was performed for the samples with 0.6x and 0.2x DNA easy clean beads. Quality check for the single stranded circular libraries was performed using Qubit SS DNA kit. Library concentrations in the range of 0.6-3 ng/ul were considered qualified for DNA Nanoball (DNB) preparation. Samples with DNB concentrations between 8ng/ul to 40ng/ul were pooled and loaded onto to the DNBSEQ T10 flowcell and sequenced using DNBSEQ-T10RS DNB Sequencing Set (FCL PE100) (940-000078-00, MGI, Shenzhen, China). The recorded data was analysed using ZLIMS Elite v1.0.5.2 software with MEGABOLT_2 pipeline.

#### ONT

Library preparation was carried out using Ligation sequencing kit 114. A total of 1000 ng DNA in 50ul was used for library preparation. Preparation steps included normalization, mechanical fragmentation using FastPrep, end repair, adapter ligation. Quality check for the double stranded libraries were performed using Qubit ds DNA kit. Libraries with 400ng/ul were loaded onto to the PromethION flowcell and sequenced using PromethION 48. The recorded data was analysed using MinKNOW software with Dorado.

### Analysis pipeline for each HTS platform

#### Illumina

The BCL file is the native output format of Illumina sequencing systems. We used the on-premise Illumina DRAGEN germline pipeline host software version 4.1.7 (DRAGEN Bio-IT Platform developed by Illumina) (Behera et al., 2024) to de-multiplex and base-call BCL files into per-sample FASTQ files. DRAGEN germline pipeline accelerates the secondary analysis of NGS data. For example, the time taken to process an entire human genome variant calling at 30x takes around 20 minutes. DRAGEN is used to preprocess the FASTQ files (adapter trimming, quality filtering) and the resulting reads are aligned to the GRCh38 human reference genome. Further, post-alignment the identification of various genetic variations such as single nucleotide polymorphisms, insertions and deletions, copy-number variations (CNVs), single tandem repeats (STRs), human leukocyte alleles (HLA) are performed by the DRAGEN variant caller with high accuracy and speed. The alignments and genetic variant calls are stored in CRAM and VCF/gVCF format respectively for any tertiary analysis. Summary statistics of alignment/variant calls and various logs of tools from DRAGEN are stored.

#### MGI

The CAL file is the native output of MGI sequencing systems. We used the on-premise MGI Ztron Pro to de-multiplex and base-call CAL files into per-sample FASTQ files. MGI’s FPGA-based hardware acceleration, ZBOLT Pro is a rack server that provides higher analysis capacity to perform bioinformatics analysis on data generated from MGI’s sequencers. ZBOLT Pro was used to preprocess the FASTQ files (adapter trimming, quality filtering) and the reads are aligned to the GRCh38 human reference genome. After mapping to the host genome, the mutation detection is performed by ZTRON Pro which yields genetic variations such as single nucleotide polymorphisms, insertions and deletions. The alignments and genetic variant calls are stored in CRAM and VCF/gVCF format respectively for any tertiary analysis. Custom in-house pipeline on G42 Cloud was used for performing the analysis of copy-number variations (CNVs using CANVAS, v1.40.0.1613) (Roller et al., 2016), single tandem repeats (STRs using Expansion Hunter, v5.0.0) (Dolzhenko et al., 2017), human leukocyte alleles (HLA using HLA-LA, v1.0.3) (Dilthey et al., 2019) using the CRAM and VCF outputs from ZBOLT. Summary statistics of alignment/variant calls and various logs of tools from ZBOLT are stored.

#### ONT

FAST5 file is the native output of PromethION (P48) sequencing system. Each P48 is tagged with an NVIDIA A100 Tensor GPU (https://www.nvidia.com/en-us/data-center/a100/) to de-multiplex and base-call FAST5 files into per-sample FASTQ/uBAM files. G42 clouds hosts an in-house custom pipeline including ONT-recommended tools for processing the uBAM files. Multiple uBAM files are merged into a single uBAM file per sample using Samtools, v1.19 (Danecek et al., 2021). On each sample, Fastp (v0.23.4) (S. Chen et al., 2018) is performed for initial sequencing QC and check if the target total number of Gb was achieved during the sequencing. We aligned uBAM file to the GRCh38 human reference genome using Sentieon’s acceleration of Minimap2 (v2.22) (Li, 2018) and we used Alfred (v0.2.6) (Rausch et al., 2019) to check the quality and alignment QC for each sample. The alignments generated by Minimap2 were stored in CRAM format. We used the alignments to call SNV, INDELs and SV using Clair3 v1.0.4 (Zheng et al., 2022) and Sniffles2 v2.2 (Smolka et al., 2024), respectively. Copy-number variations (CNVs using Spectre v0.2.1-alpha), single tandem repeats (STRs using Straglr v0.2.4) (Chiu et al., 2021) human leukocyte alleles (HLA using HLA-LA v1.0.3) (Dilthey et al., 2019) and survival motor neutron (SMN1/2) using Hapdup (v0.12.3) / Hapdiff (v0.8.8) (Kolmogorov et al., 2023) were also part the in-house pipeline. We used VariantQC (Yan et al., 2019) for performing the quality checks on the variants called and reporting the statistics for each sample. We primarily kept alignments (in CRAM format) and genetic variants (VCF and / or gVCF) as well as software tools logs and summary statistics.

### PCA

Beyond the standard QC metrics, PCA emerges as a powerful tool for uncovering underlying patterns and discrepancies in sequencing data. This step presents an additional layer of QC using PCA to assess the consistency across samples sequenced on multiple platforms, including Illumina, MGI, ONT R9, and ONT R10. PCA is a statistical procedure that transforms a set of observations of possibly correlated variables into a set of values of linearly uncorrelated variables called principal components. The transformation is defined in such a way that the first principal component has the largest possible variance, and each following component is structured to capture the maximum variance possible, provided it remains orthogonal to those before it. In this study, PCA was applied as an additional QC measure to assess the consistency of sequencing data across different platforms: Illumina, MGI and ONT (R9 and R10). Seventeen Coriell samples were sequenced on each of these platforms, generating a comprehensive dataset for analysis. After the analysis, the two main components of this PCA were visually summarized in **Supplemental Figure S5**.

### SNV/INDEL variant calling performance evaluation

Evaluating the performance of variant calling tools is crucial in genomics research and clinical diagnostics to ensure the accuracy and reliability of genetic variant identification across the whole genome. This evaluation process involves comparing the variants identified by our variant caller (which are known as test variants) against a known set of variants (reference or “golden truth”) for a given set of samples. The metrics commonly used for this assessment are recall, precision, and F-score.

#### Recall (or sensitivity)

Recall measures the variant caller’s ability to correctly identify variants present in the golden truth dataset. It is calculated as the number of true positive variants detected divided by the sum of true positive and false negative variants in the reference set.

#### Precision

Precision assesses the proportion of identified variants that are true positives. It is determined by dividing the number of true positive variants by the total number of variants called (the sum of true positives and false positives).

#### F-score

F-score is a harmonized metric that combines recall and precision into a single value, providing a balanced measure of the variant caller’s overall performance.

To evaluate the variant calling performance of SNVs and INDELs, we analyzed samples GM24149, GM24143, and GM24385, which were sequenced on each of the presented HTS platforms. The resulting VCF from variant calling was compared to its golden truth, filtered according to whether the performance was to be assessed genome-wide or at the CMRG level. Genome-wide variant calling performance was evaluated using Illumina’s hap.py script (https://github.com/Illumina/hap.py), while assessments at CMRG level employed the Real Time Genomics (RTG) tool (https://github.com/RealTimeGenomics/rtg-tools). Both tools provided detailed metrics including the number of false positives (FP), true positives (TP), and false negatives (FN).

For the SV analysis, sample GM24385 was used in Illumina, ONTR9, and ONTR10. MGI was not utilized as its pipeline does not include structural variant calling. In this case, the comparison between the generated VCFs and the golden truth, both genome-wide and at the CMRG level, was performed using the bench function of Truvari.

### Pairwise genotype concordance between platforms

We measured genotype concordance between sequencing platforms using the Jaccard index, a statistical measure for comparing the similarity and diversity of sample sets. The Jaccard index was calculated from the variant call sets in pairs of sequencing samples from the same Coriell specimen. This provided a quantitative measure of the proportion of shared variants relative to the total number of unique variants, thereby reflecting the similarity and concordance of the variant call sets across platforms.

To ensure high-confidence data, variants that passed all quality control filters from the VCF were first retained. These filtered variants were then categorized into single nucleotide variants SNVs and INDELs to facilitate independent analysis of each type. The Jaccard index was subsequently computed for the specified pair-sets using BEDTools.

## Data Availability

All data availability supporting the findings of this study may be discussed with the corresponding author upon reasonable request

## ACKNOWLEDGEMENTS

This study used samples (GM04099, GM10684, GM07828, GM07829, GM07830, GM08211, GM10354, GM13708, GM27630, GM27631, GM27632, GM03620, GM03990, GM09145, GM24143, GM24149, GM24385) from the Coriell Institute for Medical Research.

Whoever from M42 who is not author.

## AUTHORS CONTRIBUTIONS

<u>Sponsor and supervision</u>: Albarah El-Khani (AEK), Tiago Magalhaes (TM), Val Zvereff (VZ), FJS, and Javier Quilez (JQ). <u>Study design</u>: Santosh Elavalli (SE), Shalini Behl (SB), TM, VZ and JQ. <u>Data</u> <u>generation</u>: SB, Fatima Aldhuhoori (FA), Thyago Cardoso (TC), Joseph Mafofo (JM). <u>Data pre-</u> <u>processing</u>: SE, Abdelrahman Ahmed Yehia Abdelaziz Saad (AS), Gurunath Katagi (GK), Omar Soliman (OS), Shilp Purohit (SP), Ayesha Al Ali (AA), Vinay Kusuma (VK) and Haiguo Wu (HW). <u>Data</u> <u>analysis</u>: Judith Arres (JA), SE, Daniel Sanchez (DS), Luis F Paulin (LFP), JQ and Philippe Sanio (PS). <u>Manuscript writing</u>: JA, SE, DS, SB, FJS and JQ. <u>Critical feedback and manuscript revision</u>: All authors.

