## Supplemental Material for "Assessing the readiness of Oxford Nanopore sequencing for clinical genomics applications"

### SUPPLEMENTAL FIGURES

#### Supplemental Figure S1. ONT sequencing data analysis workflow.

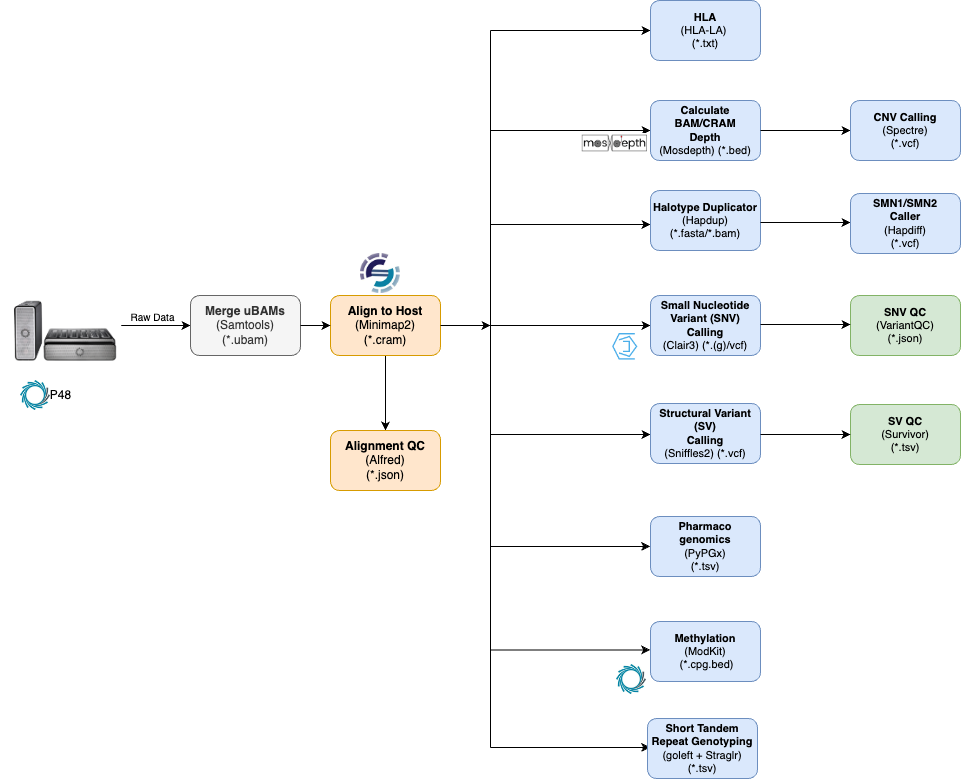

#### Supplemental Figure S2. Sequencing yield and mapping mean coverage distribution across samples and platforms.

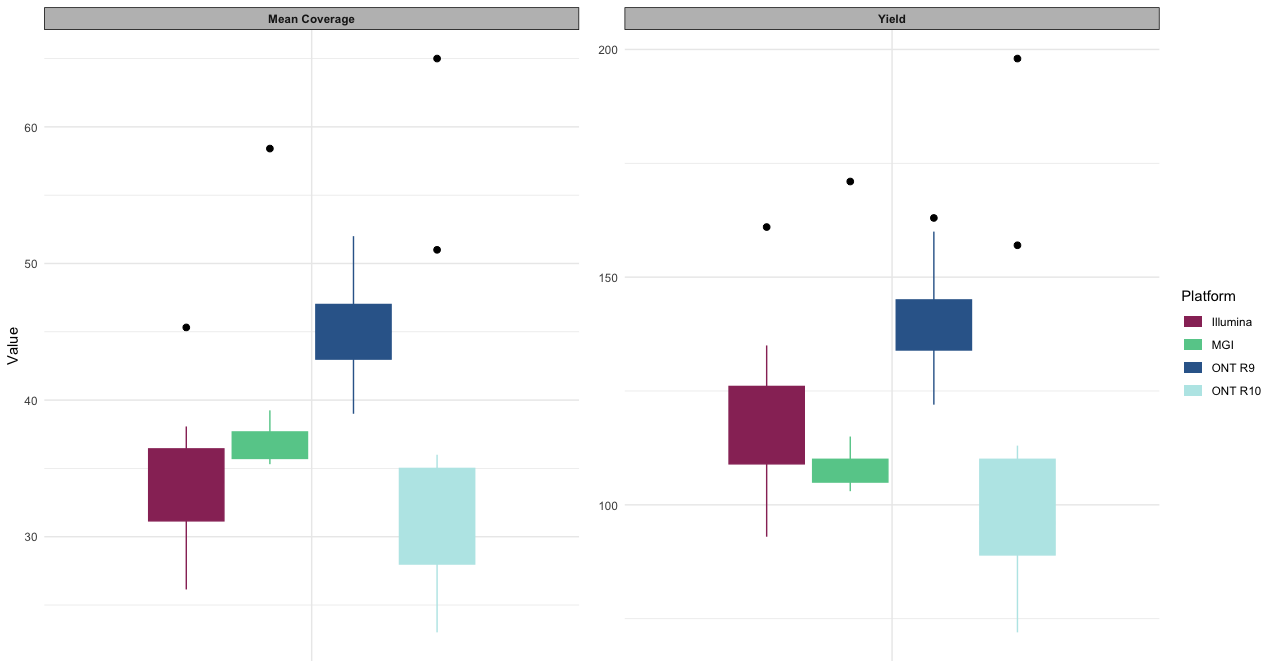

#### Supplemental Figure S3. Read quality distribution in ONT samples.

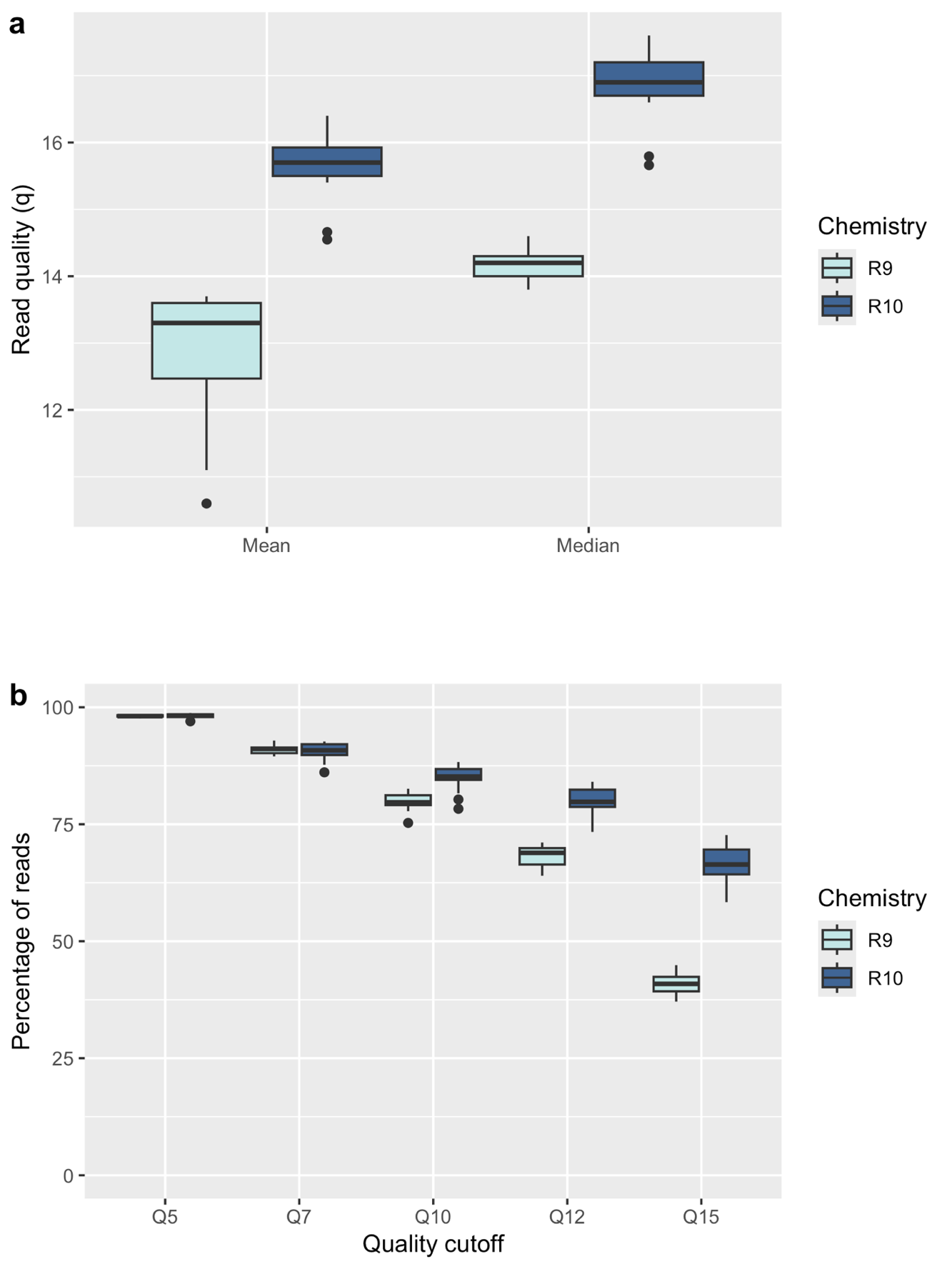

#### Supplemental Figure S4. Read length distribution in ONT samples.

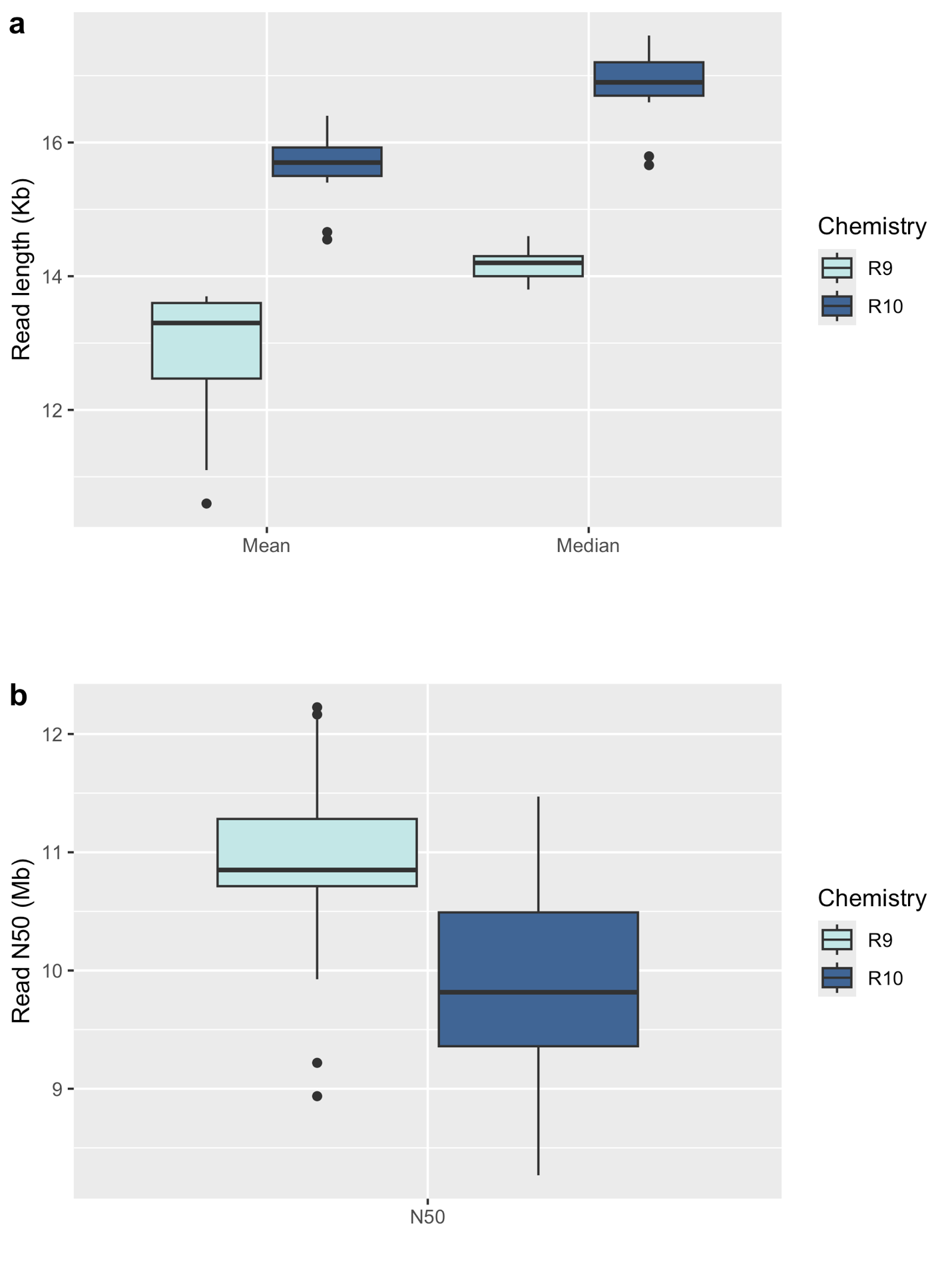

#### Supplemental Figure S5. Number of small variants called per sample across the four sequencing platforms.

The boxplot shows the distribution of SNVs and INDELs identified in Coriell samples sequenced using Illumina (4.0M SNVs, 1.0M INDELs), MGI (4.0M SNVs, 0.9M INDELs), ONT R9 (4.5M SNVs, 1.0M INDELs), and ONT R10 (4.5M SNVs, 1.2M INDELs).

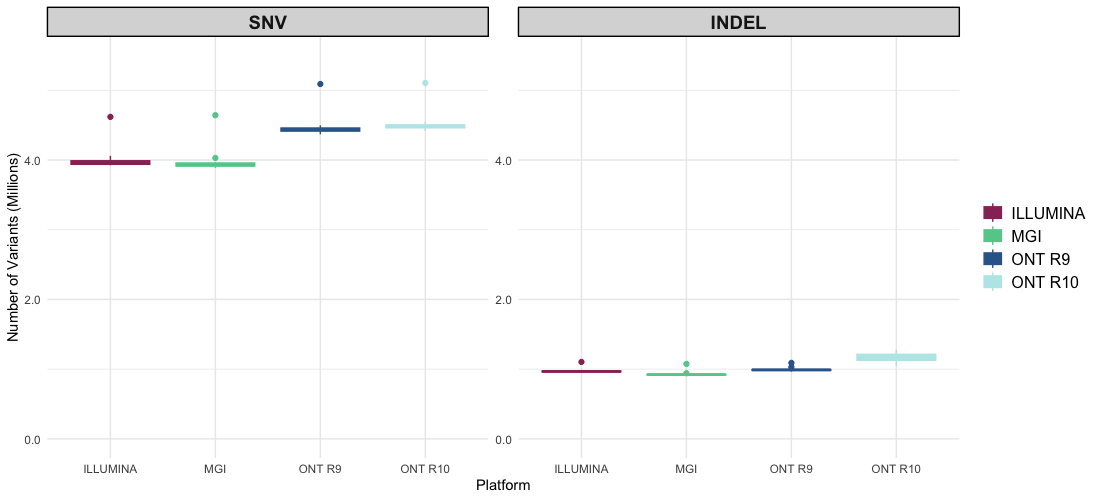

#### Supplemental Figure S6. Clustering of the 17 Coriell reference samples reflects ancestry and familiar relationship.

**(a)** Scatter plot of PC1 versus PC2 following PCA on the SNP/INDEL calls for the 17 Coriell reference samples. We manually labelled the main three ancestries (African, Caucasian and South American). **(b)** Same plot as in (a) but removing the samples of African ancestry, which makes more visible that samples from the same individual cluster together and the two families do it too. South American trio: GM27630 (son), GM27631 (father) and GM27632 (mother). Caucasian family: GM24143 (mother), GM24149 (father) and GM24385 (son).

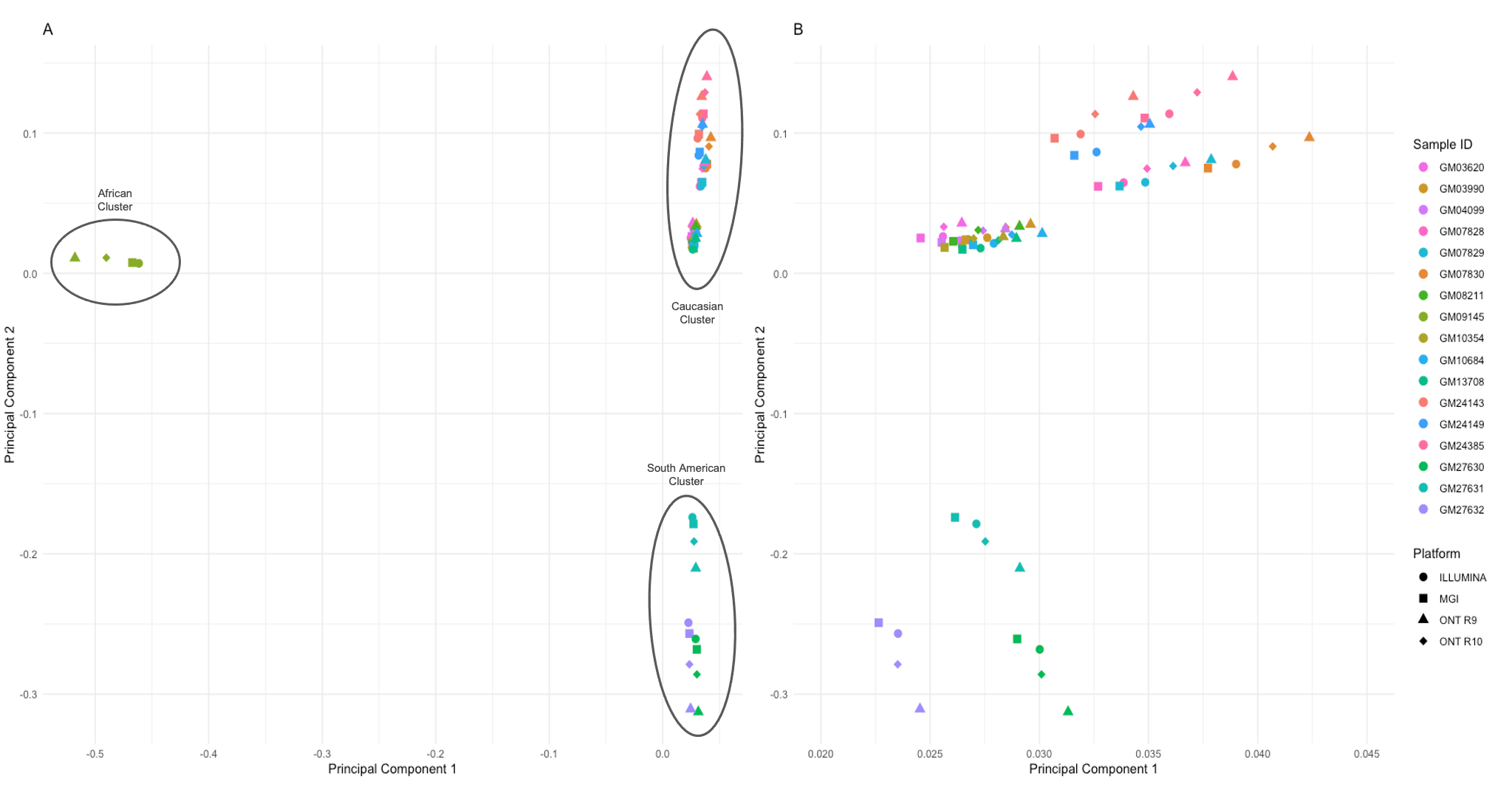

#### Supplemental Figure S7. PC4 captures differentiates sequencing technology.

Boxplots of the first four PCs from the PCA, grouped by sequencing platform (Illumina, MGI, ONT R9, ONT R10).

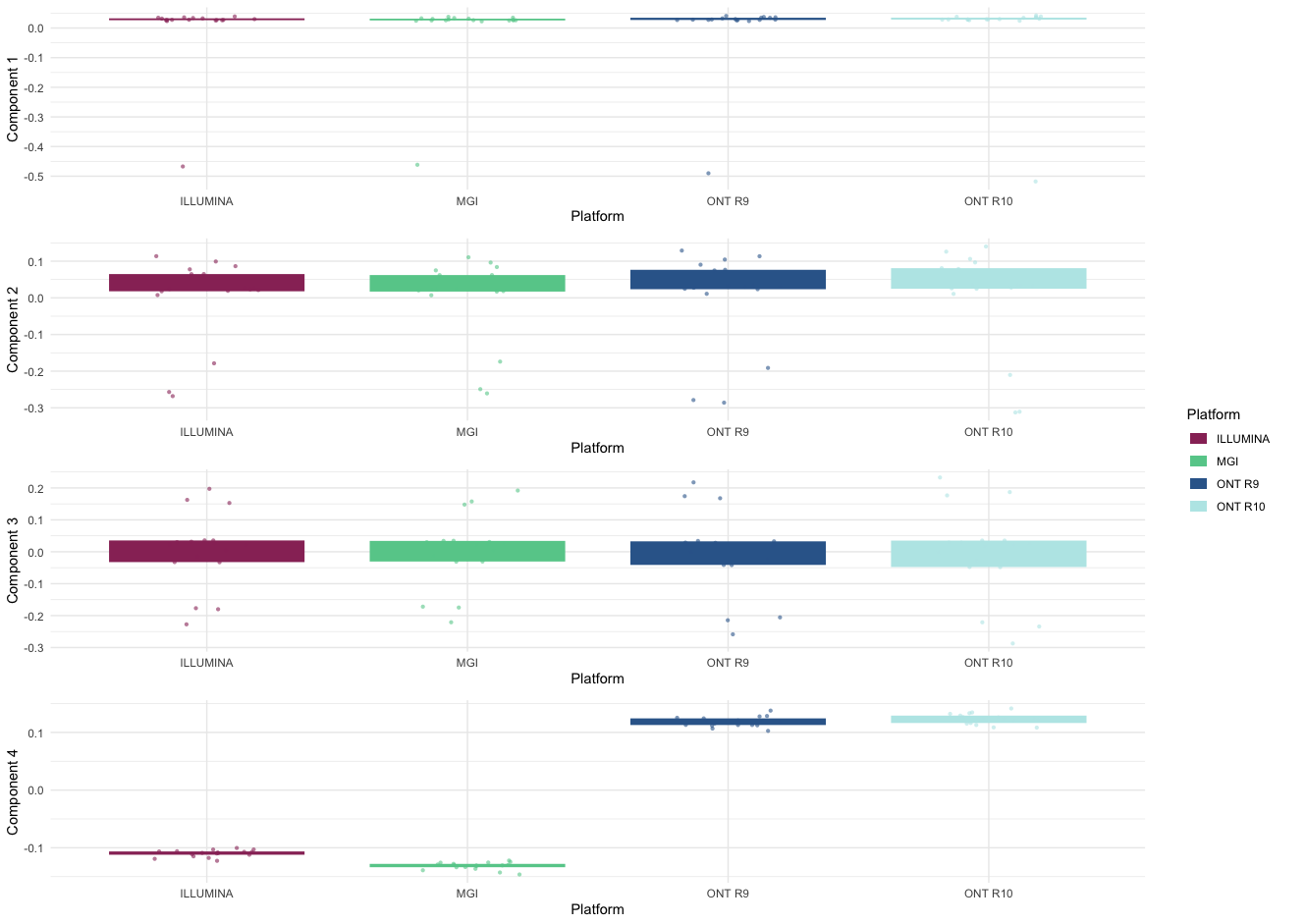

#### Supplemental Figure S8. SNV variant calling performance.

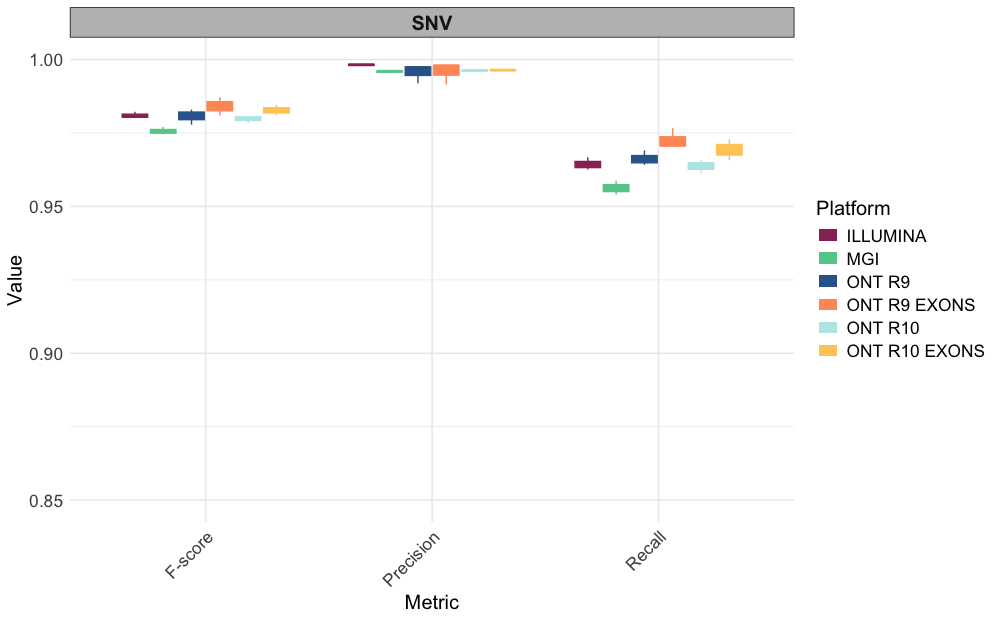

#### Supplemental Figure S9. SNV and INDEL variant calling performance in CMRG for GM24385.

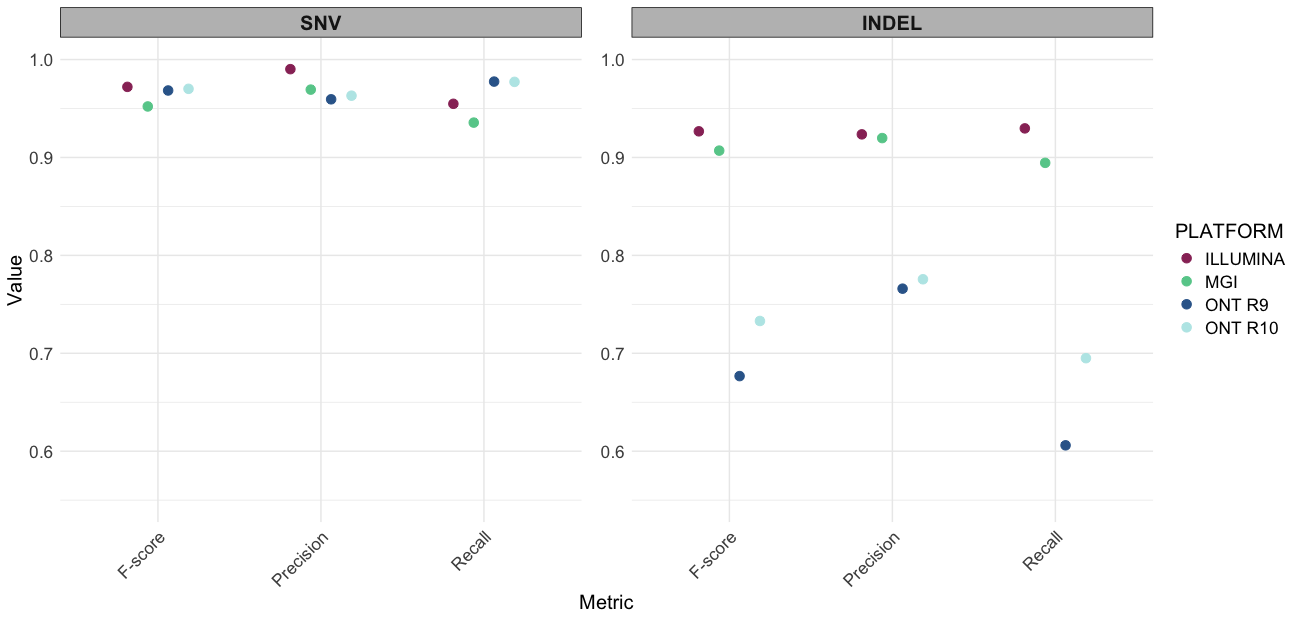

#### Supplemental Figure S10. IGV screenshots for chromosome X in the region of the *DMD* gene for sample GM04099.

(A) ONT R9 results and in (B) ONT R10 results. In both cases, the decrease in coverage denoted by the smaller pile-up of aligned reads hints the presence of the deletion.

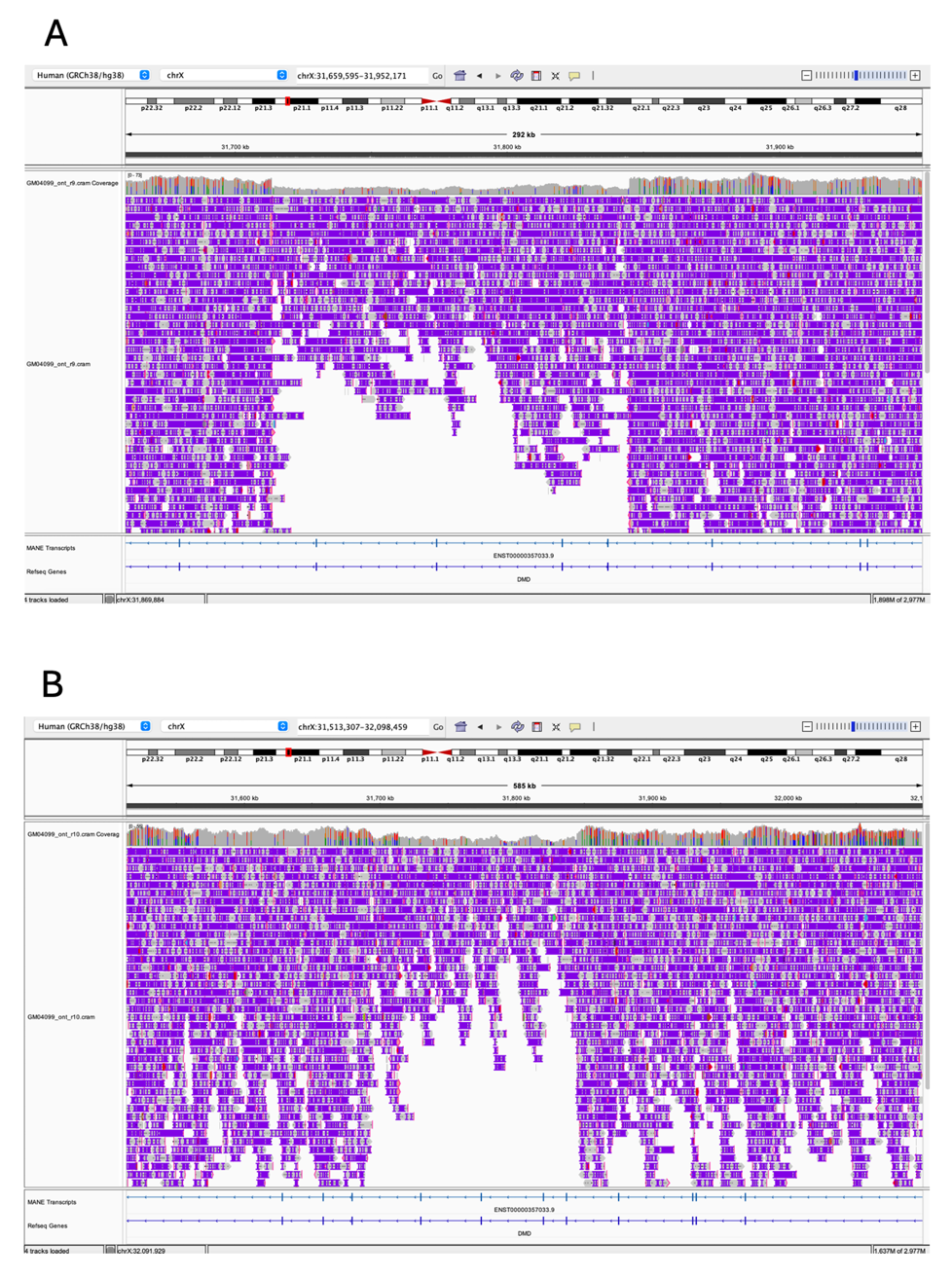

#### Supplemental Figure S11. SNV density chromosome profiles across platforms.

The dashed black lines mark the centromeres and, for chromosome 6 only, the dashed red lines indicate the HLA region.

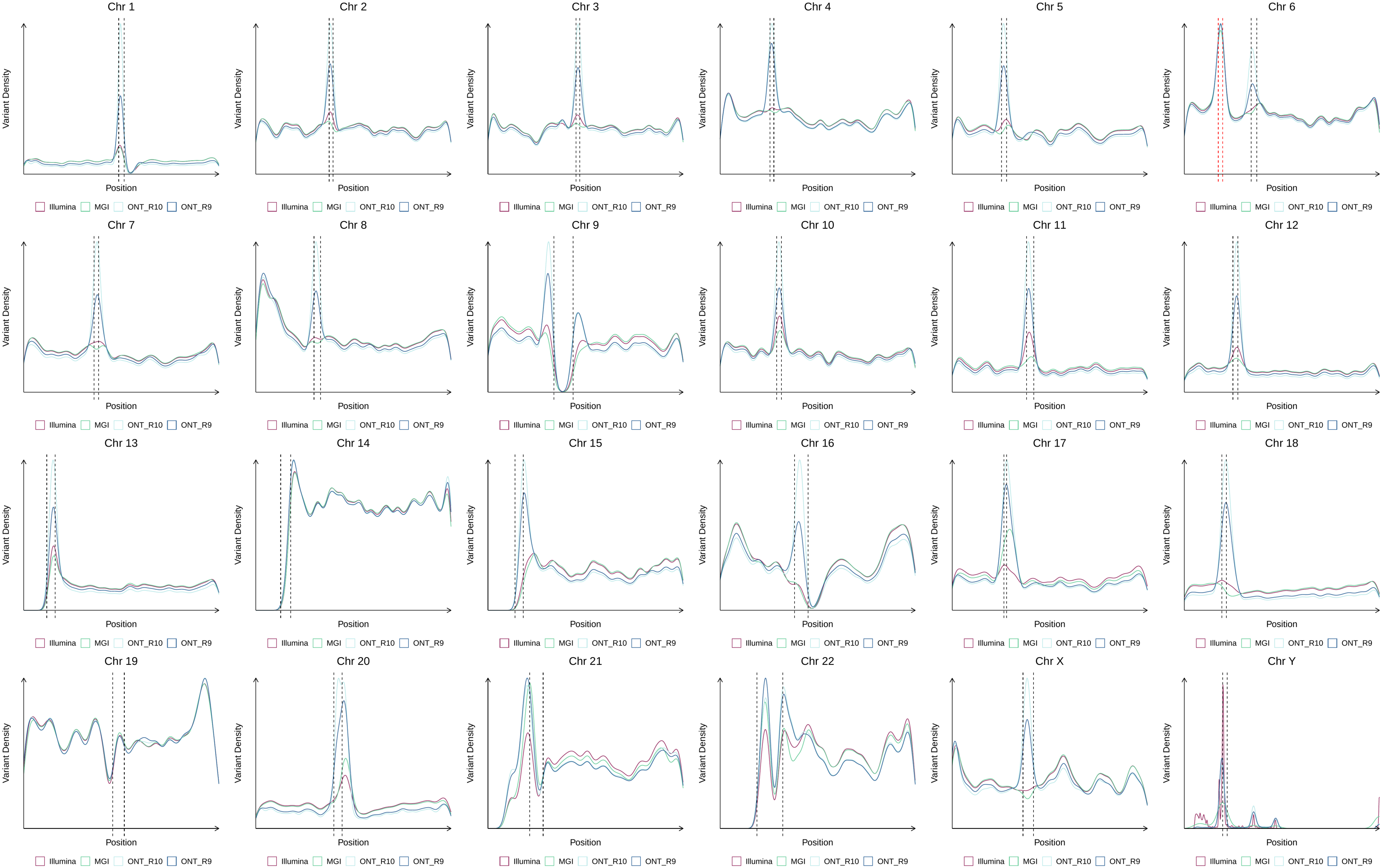

#### Supplemental Figure S12. SNV and INDEL genotype concordance between sequencing platforms.

The Jaccard index values were calculated to assess the concordance of SNV and INDEL detection across the different sequencing platforms. SNVs display consistently high concordance, indicating reliable detection capabilities across platforms. In contrast, INDELs show significantly lower concordance, particularly in the comparison between short and long reads platforms.

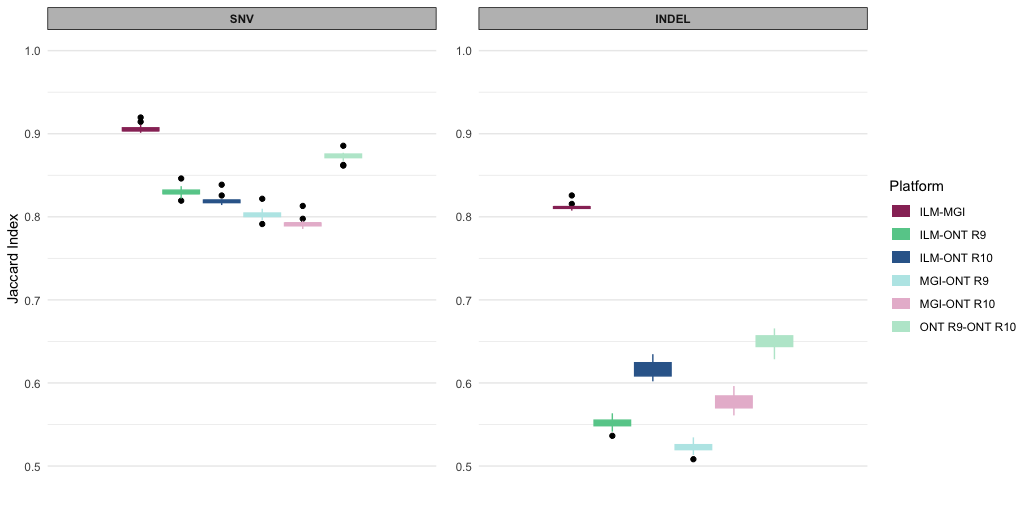

### SUPPLEMENTAL TABLES

#### Supplemental Table S1. QC metrics and high-quality cutoffs to filter WGS datasets

#### Supplemental Table S2. 17 Coriell reference samples used in this study

Highlighted in grey are the 3 samples used to determine genome-wide performance metrics instead of detection of point disease-causing mutations.

#### Supplemental Table S3. Summary of QC metrics across the 76-samples WGS dataset.

Yield is the total amount of DNA sequenced expressed in gigabases (Gb). The percentage of bases >Q30 and genome >10X are self-explanatory. Mapping rate is the percentage of sequenced bases that align to the reference genome. Mean coverage is the number of times a genomic region has been sequenced on average. Het/hom ratio consists of balance between heterozygous and homozygous variants (which is a proxy to detect possible contamination).

#### Supplemental Table S4. Performance calling SV (a) genome-wide and in (b) CMRG for GM24385.

#### Supplemental Table S5. Genotyping results for the 11 evaluated pathogenic mutations across all 14 Coriell positive controls and all 4 sequencing platforms.

Each cell in the table shows the genotype for each variant (column) for each sample (row).

Highlighted in green and red, respectively, are the successfully (true positive) and inaccurate (false negatives) genotyped pathogenic variants. Blank denotes genotypes in which the pathogenic variant is absent (true negatives).

For “DMD EX49-52DEL", the value “1” indicates the deletion of exons 49–52 in *DMD* and 0 no deletion.

For “SMN1 EX7-8DEL", the left and right value of the “/” indicates the number of copies of *SMN1* and *SMN2* respectively – zero copies of *SMN1* corresponding to the pathogenic deletion causing SMA. We only run Sillago SMN1/2 caller on the SMA-positive ONT R9 and R10 samples so it is absent (“-”) for the negative controls. For the STRs “DMPK CTG”, “FMR1 CGG” and “HTT CAG” we provide the STR copy number. “FMR1 CGG” is in chromosome X so a single value is reported for males.

#### Supplemental Table S6. Jaccard index values between platforms for SNV and INDEL.

### SUPPLEMENTAL NOTES

#### Supplemental Note 1. Overcoming the challenges of ONT WGS data analysis

The initial step in the processing of sequencing data generated by ONT sequencers is the base-calling. This step consists in determining the nucleotide sequence (DNA or RNA) from the electric squiggle stored in the Fast5 files produced by the sequencing instrument. Because methylation modifications in the nucleotide sequence are reflected in the squiggle, such modifications can be captured during the base-calling. We refer to canonical base-calling to only determining the nucleotide sequence and concurrent base-calling to the simultaneous inference of the nucleotide sequence and its methylation modifications.

Two critical challenges in the processing of ONT data are the large size of the Fast5 files with the raw data and demanding computing requirements to perform the base-calling. The aggregated size of Fast5 files for a single ~30X human genome is ~700 Gigabytes (GB). Besides, extracting the nucleotide sequence and methylation marks from the Fast5 files requires GPU computing power and takes 24-36 hours (about 1 to 1 and half days) even with the most advanced hardware and software combination for a ~30X genome.

Over the last three years, we have been early adopters of hardware and software improvements to overcome such challenges. First, we replaced the Nvidia GV100 towers initially connected by default to ONT’s PromethION 48 sequencer with the more powerful Nvidia A100. Second, we upgraded from Guppy version 4 to version 6 to enable concurrent base-calling, and more recently to the newer Dorado base-caller to achieve a two-fold increase in base-calling performance relative to Guppy version 6 (<https://aws.amazon.com/blogs/hpc/benchmarking-the-oxford-nanopore-technologies-basecallers-on-aws/>). We observed that Guppy version 6 on Nvidia A100 achieved real-time base-calling not for a full PromethION 48 run (48 flow-cells) – instead, it could catch up with 24 and 12 flow-cells with canonical and concurrent base-calling, respectively.

Prior to these improvements, real-time concurrent base-calling was not feasible. We had to upload large Fast5 files (~700 Gigabytes per ~30X genome) to our cloud to perform base-calling, increasing the run times and the cloud storage footprint. Besides, concurrent base-calling prior to Guppy version 6 was prohibitive in terms of runtime and associated computing cost.

By default, concurrent base-calling will result in three large files (Fast5, FASTQ and uBAM) that add up to >1 TB for a ~30X human genome. The uBAM contains the same information as the FASTQ plus the methylation marks, so the latter becomes redundant. Likewise, once the base-calling completes successfully, it is not cost-effective to keep Fast5. Even if better base-callers are developed (e.g. more accurate, capturing additional methylation marks), the cost of storing the Fast5 files is likely to exceed that of re-sequencing. Therefore, we recommend to only keep the uBAM for downstream processing and to delete the Fast5 and FASTQ files (indeed the generation of the latter can be turned off to start with).

We accelerated the mapping of the ONT sequencing reads to the genome refence sequence by using Sentieon’s Minimap2 version, which is approximately two times faster than the open-source version of the mapper. Indeed, that is one of the principal caveats we would see in using ONT’s Human variation workflow, which uses the open-source Minimap2. Regardless of the mapper, we recommend storing the alignments in the CRAM (2) format (instead of BAM format). We found that, for ONT data, CRAM achieved on average ~40% compression relative to BAM, as well as data lossless and unaltered variant calls (3). In case of short nucleotide variant (SNV) calling, we have benchmarked the tool (https://github.com/HKU-BAL/Clair3) from version 1 to the current version 3 which showed significant improvements in terms of precision/recall as well the computational time improved with more newer releases. However, this SNV calling is a pain point in terms of the time taken to run a sample on gVCF mode (~ 20-24 hours for a ~30x genome) and is much faster with VCF mode (~5-6 hours). In the case of structural variant calling (SV), we used Sniffles2 that produces structural variant calls within an hour and is highly recommended.
